# Cannabis Use Documentation within the Electronic Health Record: A Use Case for Natural Language Processing Methods

**DOI:** 10.64898/2026.02.27.26347207

**Authors:** Apoorva M. Pradhan, Vishal A. Shetty, Christina M. Gregor, Jove H. Graham, Lorraine Tusing, Annemarie G. Hirsch, Eric Hall, Vanessa Troiani, Mellar P. Davis, Donielle L. Beiler, Katrina M. Romagnoli, Chadd K. Kraus, Brian J. Piper, Eric A. Wright

## Abstract

**Introduction:** Recreational and medical cannabis use (CU) information is often available within the electronic health record (EHR) in a format that is impractical for health care provider use. Transformation of free-text EHR documentation in notes to discrete elements is possible using natural language processing (NLP) and has the potential to characterize CU efficiently. The objective of this study was to develop an NLP algorithm to identify documentation of CU within EHR unstructured clinical notes.

**Methods:** We identified EHR notes with cannabis-related terminologies through a keyword search among all Geisinger patients with at least one encounter between 1/1/2013 and 6/30/2022. We trained four NLP models to classify notes into six categories based on time, context, and reliability of CU documentation identified through manual annotation. We compared the demographic characteristics of patients with positive classification for CU using the best-performing model to those of the overall population.

**Results:** Of the over 1.7 million eligible patients, 150,726 (8.6%) were flagged as cannabis users. The Bio-ClinicalBERT, a transformer-based NLP model, achieved close to human performance in classifying CU (weighted Precision=91.4, Recall=93.3, F-score=92.4). Cannabis users had higher BMI and were at least nine-fold more likely to use tobacco, alcohol, and illicit substances.

**Conclusion:** Our study evaluated the prevalence of CU documentation across the entire corpus of EHR notes data without population segmentation. The NLP methodologies used achieved performance close to that of human annotation and laid the foundation for identifying and classifying CU within unstructured data sources, with future applications in research and patient care.

**Plain Language Summary:** Marijuana, also known as cannabis, may impact the health of patients, yet it is not routinely captured in medical records, and when documented, it is often found in unstructured formats (e.g., progress notes) rather than in discrete fields. Incomplete and unstructured capture limits many functional capabilities within the EHR that enhance patient care (e.g., drug interactions, notifications) and limit researchers from identifying patients routinely exposed to marijuana use. The transformation of free-text documentation of cannabis use (CU) into discrete elements can be performed using natural language processing (NLP). The objective of this study was to develop an NLP model to identify CU in unstructured clinical notes in the EHR. We examined the EHRs of Geisinger patients in Pennsylvania over a 10-year period. Among 1.7 million patients, 9% were identified as CU. One of the NLP models tested, Bio-ClinicalBERT, achieved the highest performance. Cannabis users had a higher BMI and were ten-fold more likely to be tobacco users, ten-fold more likely to use alcohol, and nine-fold more likely to use illicit substances. NLP can be used to better understand the risks and benefits of CU at a population level and may improve patient identification to assist clinical decision-making. Future CU epidemiological research should continue to explore other avenues to automate and improve CU documentation by leveraging rapidly evolving technologies, such as artificial intelligence-driven tools.

## Introduction

Cannabis products, both recreational and medical, are commonly used in the United States (US)[1–3]. The 2024 National Household Survey on Drug Use and Health estimated that 44.3 million people, age ≥ 12, had past-month marijuana use[4]. The Commonwealth of Pennsylvania (PA) passed its Medical Marijuana (MMJ) Act in 2016[5]. As of November 2025, more than one out of every twenty-five PA (PA) adults (3.37%) were certified for medical marijuana[6] with anxiety disorders, chronic pain, and post-traumatic stress disorder accounting for the preponderance (85.6%) of certifying conditions (Supplemental Figure 1)[7,8].

The medical marijuana certification process in PA, like most other states, is typically completed by a physician with whom the patient does not have a long-standing relationship, as they have with their primary care provider, often by telemedicine [9,10]. Three-quarters of states, including PA, that have a medical marijuana program do not report any information to their Prescription Drug Monitoring Program [11]. With the executive order from December 2025 facilitating the potential reclassification of cannabis from a Schedule I to a Schedule III substance, there are a multitude of reasons why physicians and pharmacists need readily available CU information. These include recognition of cannabis-drug interactions [12,13], alignment with evidence driven utilization for medical conditions for which CU is indicated [14–16], awareness of CU as a substitute for prescription medications [17], aiding diagnosis of CU-related conditions (e.g. cannabis use disorder, hyperemesis syndrome, etc.) [18], and awareness of CU can assist clinicians in discussing safe use practices (e.g. storage in homes with children) [19]. The examples listed above highlight some of the scenarios that explain the necessity behind improving the understanding of an individual’s cannabis use and its importance in making informed health decisions.

Despite the need, the amount of actionable information documented in electronic health records (EHR) remains inadequate (e.g., only 4.8% of medical CU is documented in the EHR compared to 35.1% that is implicitly reported)[20,21]. The most common source of CU documentation is unstructured clinical notes [22–31] or patient portal messages [32]. A prior report from members of this team in PA discovered that secure patient portal messages between patients and health care providers discussing cannabis from 2012-2022 peaked in 2019 after dispensaries opened in 2018 [32]. Natural language processing (NLP) models have been leveraged to identify CU-related documentation from unstructured text [26–31]. Most prior studies applying NLP for CU identification have focused on specific sub-populations by disease states, such as psychosis patients, primary care patients, older adults undergoing surgery [26–28,30,31,33] or limited age ranges like pediatric and young adults, [26,29,31], thereby limiting their applicability for understanding CU across the entire population captured within the EHR. The purpose of this investigation was to extend prior studies [25,32] and to use validated NLP methodologies to classify CU from unstructured EHR notes, providing a proof of concept that free-text EHR notes can be extracted for CU using an automated mechanism from a broad, unstratified population [26].

## Methods

We adhered to the STROBE reporting guidelines for reporting purposes (Supplement F, [34]). Figure 1 presents the pipeline and phases involved in this study.

**Fig. 1.**
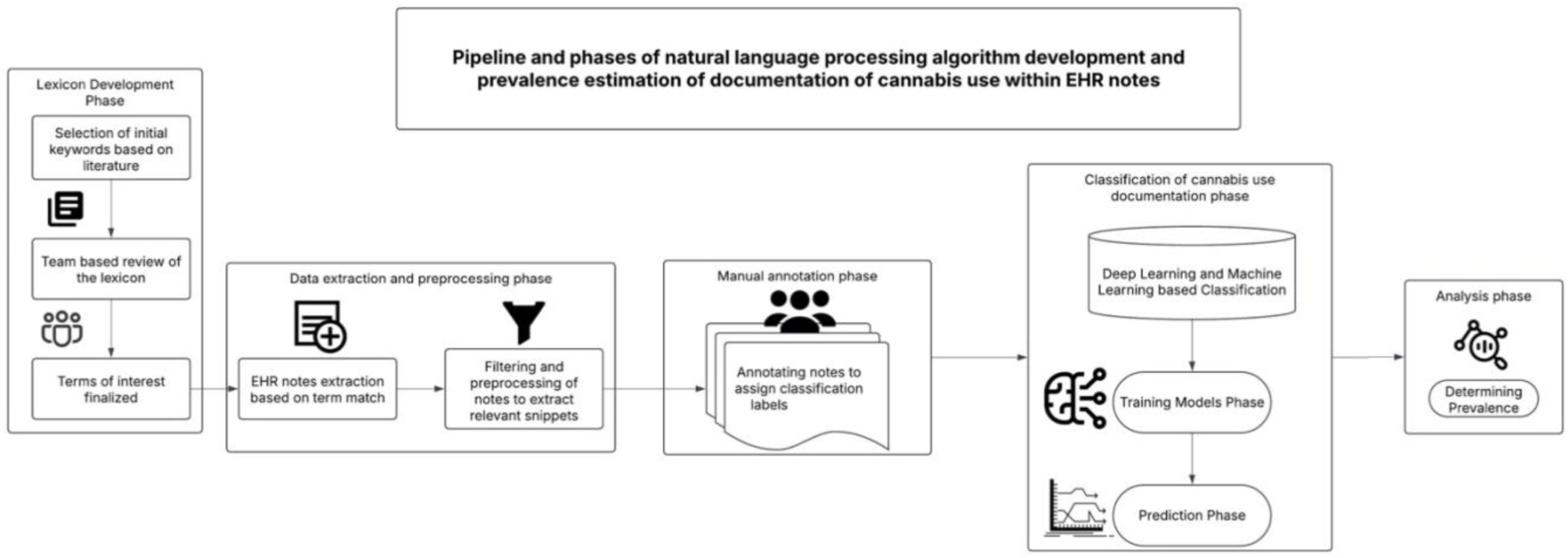
Diagram of the pipeline and phases involved in this study for the development of a natural language processing algorithm to assess the prevalence of cannabis use documentation over a period of 9.5 years within electronic health record notes.

### Study Setting

This investigation was conducted at Geisinger, an integrated health system in the US, spanning 24 counties in central and northeast PA [Supplemental Figure 2A]. Geisinger comprises ten acute care hospitals, 133 primary and specialty clinics, a research institute, and an insurance company, Geisinger Health Plan. Geisinger began implementing its EHR, Epic^®^ Corporation (Verona, WI), in 1996 at all ambulatory sites and integrated it across inpatient care sites in 2003. This infrastructure provides a comprehensive view, including clinical information, imaging, and administrative claims for rural and urban residents, offering a real-world examination of dynamic changes in this population.

### Patient Population

The study included patients with at least one encounter (e.g., inpatient, outpatient, emergency department) at a Geisinger facility between January 1, 2013, and June 30, 2022. This population will henceforth be referred to as the overall population. For patients included in the overall population, we only pulled EHR notes that included one of the cannabis-related terms from the lexicon. Although population declines occurred in 20 of 24 (83.3%) counties (Supplemental Figure 2B), nativity levels remained high in the Geisinger footprint (Supplemental Figure 2C), supporting the use of this population for the extended period employed in this investigation.

### Lexicon Development and Dataset Construction

This report used snippets of encounter notes curated from the EHR, including nursing notes, appointment notes, problem list-based free-text documentation, visit reason documentation, and follow-up notes. (Supplement A – Data sources)

We developed a CU lexicon to narrow the scope of data retrieval. The CU-related terms in the lexicon were finalized based on prior literature [26,27] and input from the study team’s subject matter experts (MPD, CKK, BJP). The lexicon included the following terms: ‘marijuana,’ ‘cannabis,’ ‘MJ,’ ‘THC,’ ‘CBD,’ ‘weed,’ ‘MMJ,’ ‘indica,’ ‘sativa,’ ‘cannabinoid,’ ‘spice,’ ‘tetrahydrocannabinol,’ ‘pot,’ ‘cannabidiol,’ ‘ganja,’ ‘grass,’ ‘hash,’ ‘hashish,’ ‘bong,’ ‘Mary Jane,’ ‘edibles,’, and ‘joint.’ Among the 1.76 million patients seen at Geisinger during the study period, we identified 1.71 million patients (135,858,323 EHR notes) with exact-word matches in their EHR notes using this lexicon. We then trimmed these EHR notes to retain only 300 characters before and after the CU-related term. We will refer to these trimmed snippets of notes simply as EHR notes in the context of this study moving forward. We observed that specific terms were used in contexts unrelated to cannabis; for instance, the term ‘pot’ was used in the context of ‘neti pot,’ ‘netti pot’, or as a part of the word ‘hypotension,’ ‘potential,’ or ‘potassium,’ while the term ‘CBD’ was used in an anatomical context for the ‘Common Bile Duct’ such as “CBD measure” and “CBD stricture”, these phrases and words were removed while data cleaning. ‘CBD’ or ‘pot’, only when mentioned in the context of marijuana use, was retained in the final analytical sample for classification. The term ‘joint’ was also primarily and extensively used in an anatomical context, such as “joint pain,” “joint space,” “joint deformity,” and “joint disease”. To minimize such misclassifications, we removed EHR notes that contained these specific phrases or words (Supplement A – Primary Data Cleaning), resulting in a working sample of 17,789,648 EHR notes. The data cleaning process also involved the removal of notes when the term of interest was negated (e.g., “no cannabidiol use” or “tetrahydrocannabinol (THC): no.”) This led to a more concise final list of terms and a final analytical sample of 2,790,896 EHR notes with at least one mention of a key term in patient charts. We used SAS Enterprise Guide 8.3 for dataset construction and cleaning.

### Manual Annotation Phase

We developed an annotation schema and decision rules for labeling CU in EHR notes. VS and AP manually reviewed and double-coded a random sample of 100 EHR notes using labeling categories informed by previous literature [26]. Labeling categories were refined following a discussion of the assigned codes, with further input from team members to resolve disagreements. (Supplement B – Labeling Guidelines) This review phase eventually led to the creation of six categories:

1. True mention – The identified key term in the EHR notes refers to cannabis
2. Patient use – The true mention applies to the patient’s use, not another person
3. Indication of use – The EHR note explicitly mentions or indicates that the patient has used cannabis at some point
4. Medical use – The EHR note explicitly mentions that the patient used it for a medical purpose
5. Current use – The EHR note explicitly mentions that the patient currently uses
6. History of use – The EHR note explicitly mentions that the patient has a history of use

Three annotators were trained using the decision rules constructed for each labeling category and by reviewing the 100 initially sampled EHR notes. (Supplement C – Decision Rules) Each annotator subsequently independently reviewed a new random sample of 50 EHR notes (300 potential labels) (Supplement D – Examples), and we calculated an inter-rater reliability kappa for each category between each reviewer. (Supplement E – Inter-rater reliability kappa) Annotators then labeled EHR notes independently, generating 3,650 unique training examples. Throughout this phase, annotator questions regarding uncertain labeling decisions were discussed and resolved among the study team’s experts to refine category definitions and decision rules further. The annotated sample was drawn based on cannabis-related terms in the lexicon and was not adjusted for any patient characteristics. All annotation work was conducted using Microsoft Excel™.

### Model Development and Training

We trained two traditional models and two transformer-based models to identify CU in EHR notes: (1) logistic regression (LR), (2) support vector machine (SVM), (3) Bidirectional Encoder Representations from Transformers (BERT) [35], and (4) Bio-ClinicalBERT [36]. Prediction of each CU category was treated as a separate binary classification task for each model. Model development and training were conducted using Python 3.10.13.

For the LR and SVM models, we used a bag-of-words approach using unigrams, bigrams, and trigrams to represent each EHR note. We removed stop words and built bag-of-words representations using the sci-kit learn library [37]. We used elastic net regularization for dimensionality reduction in the LR models, with an L1 ratio of 0.5, which applies both L1 and L2 penalties equally. For SVM, a linear kernel was applied. We tested regularization weights of 0.01, 0.1, 1, 10, and 100 for LR and SVM. We used a regularization weight of 0.01 for LR models and 10 for SVMs, as model performance was highest with these weights (based on f-score across all CU categories). We conducted 5-fold cross-validation to train and evaluate models, averaging performance metrics across each test fold. Precision, recall, and F-score were used to assess model performance.

For BERT and Bio-ClinicalBERT, we used each model’s pre-trained transformer layers and tokenizers available through Hugging Face’s transformer library [36,38]. Models were fine-tuned on the corpus of labeled EHR notes. The training ran for ten epochs with a batch size of 8, and the AdamW algorithm was used for optimization. We performed 5-fold cross-validation for training and evaluation, and used precision, recall, and F scores averaged across each test fold as performance metrics. Precision, or more commonly known as positive predictive value, is the proportion of positive results that are true positives in the predicted sample [39]. It answers the question, “if the patient is classified as a CU, what is the probability they actually engage in CU.” [39] Recall, also known as sensitivity or true positive rate, quantifies how well a test identifies true positives (i.e., correctly identifies subjects who truly have the condition of interest)[39]. An F-score combines precision and recall into a single score and is used to evaluate the performance of the machine learning model. All BERT and ClinicalBERT models were implemented using Pytorch 2.1.0. Our model training and testing scripts are available at [40].

### CU prevalence and patient characteristics

After evaluating each trained model, we used the highest performing model (in terms of weighted F score) to assign ‘True Mention’ and ‘Indication of Use’ classifications to all unlabeled EHR notes. Patients who had an EHR note positively classified for both the ‘True Mention’ and ‘Indication of Use’ categories were flagged as cannabis users. Any patient with at least one EHR note being flagged positive for CU was subsequently identified as a cannabis user for the remainder of the study period for descriptive analysis purposes.

We assessed the prevalence of CU documentation in the EHR using the CU flag. We computed descriptive statistics in patients flagged for CU at the time when the patient was first identified as a cannabis user, and in the overall population at the time of their first encounter during the study period. We calculated means or medians with SDs or interquartile ranges (IQRs) for continuous variables and frequencies and percentages for categorical variables, when appropriate, to evaluate the distribution of patients’ demographic and clinical characteristics including current substance use (smoking, alcohol, and illicit drug use), and BMI in the annotated sample, overall and CU populations. We further assessed the magnitude of difference in the demographic and clinical characteristics between the annotated sample, CU, and the overall population using t-tests and chi-square tests. All analyses were performed using SAS statistical software (SAS Enterprise Guide 8.3, Cary, NC).

## Results

We retrieved 2,790,896 EHR note snippets from 370,885 patients; 98.7% were captured within nursing notes. ‘Marijuana’, along with its misspelling (‘Marjiuana’), was the most used terminology (56.7%), followed by ‘CBD’ (13.3%), ‘Cannabis’ (7.9%), ‘THC’ (7.5%), and ‘Pot’ (5.1%) (Table 1). From the 3,650 EHR notes in the annotation set, 1,945 (53.3%) had a ‘True mention’ of CU. Among the ‘True mentions’, 1,911 (98.3%) were explicitly discussing the patient’s personal use at the time of the encounter. 1,476 (77.2%) had documentation about the ‘Indication of use,’ 359 (18.8%) indicated medically recommended use, 973 (50.9%) had documentation indicating ‘Current Use,’ and 551(28.8%) had a match indicating ‘Past Use.’ A high kappa (> .83) was observed between the three annotators for all the labeling categories in the training sample, except for “History of use” (Supplement C – labeling categories). Supplement Table D includes example snippets for each labeling category.

**Table 1.**
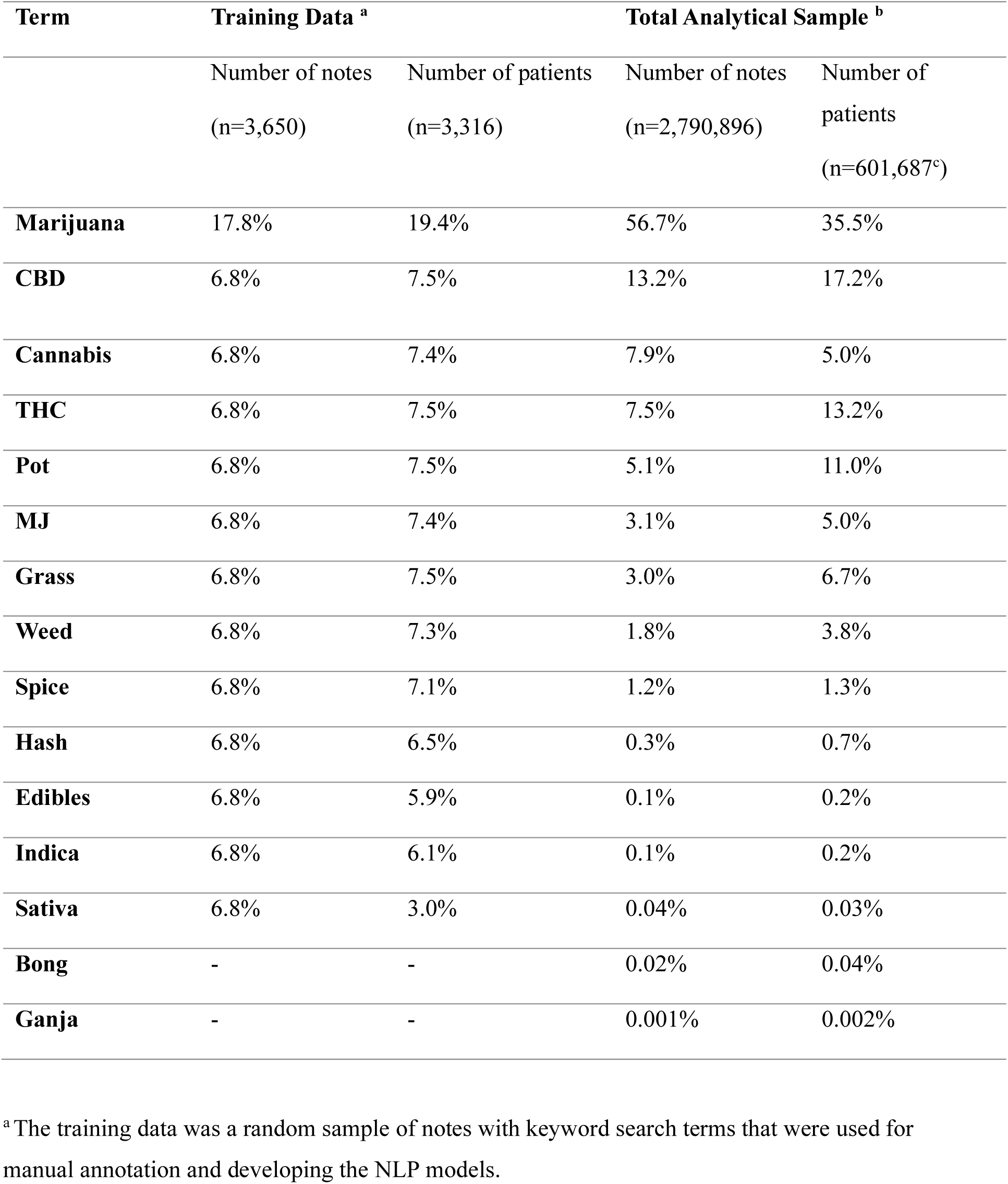
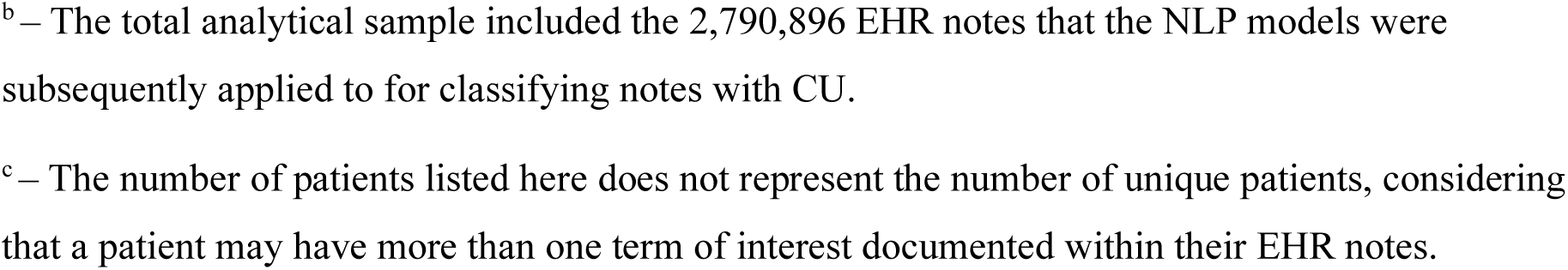
Frequency breakdown of cannabis-related keyword search terms that were used for developing classification Natural Language Processing models post data cleaning within a 2.7 million analytical sample at Geisinger over the 9.5-year study period. Cannabidiol: CBD, Tetrahydrocannabinol: THC.

| Term | Training Data <sup>a</sup> |  | Total Analytical Sample <sup>b</sup> |  |
| --- | --- | --- | --- | --- |
|  | Number of notes<br>(n=3,650) | Number of patients<br>(n=3,316) | Number of notes<br>(n=2,790,896) | Number of patients<br>(n=601,687<sup>c</sup>) |
| **Marijuana** | 17.8% | 19.4% | 56.7% | 35.5% |
| **CBD** | 6.8% | 7.5% | 13.2% | 17.2% |
| **Cannabis** | 6.8% | 7.4% | 7.9% | 5.0% |
| **THC** | 6.8% | 7.5% | 7.5% | 13.2% |
| **Pot** | 6.8% | 7.5% | 5.1% | 11.0% |
| **MJ** | 6.8% | 7.4% | 3.1% | 5.0% |
| **Grass** | 6.8% | 7.5% | 3.0% | 6.7% |
| **Weed** | 6.8% | 7.3% | 1.8% | 3.8% |
| **Spice** | 6.8% | 7.1% | 1.2% | 1.3% |
| **Hash** | 6.8% | 6.5% | 0.3% | 0.7% |
| **Edibles** | 6.8% | 5.9% | 0.1% | 0.2% |
| **Indica** | 6.8% | 6.1% | 0.1% | 0.2% |
| **Sativa** | 6.8% | 3.0% | 0.04% | 0.03% |
| **Bong** | - | - | 0.02% | 0.04% |
| **Ganja** | - | - | 0.001% | 0.002% |

<sup>a</sup> The training data was a random sample of notes with keyword search terms that were used for manual annotation and developing the NLP models.
<sup>b</sup> – The total analytical sample included the 2,790,896 EHR notes that the NLP models were subsequently applied to for classifying notes with CU.
<sup>c</sup> – The number of patients listed here does not represent the number of unique patients, considering that a patient may have more than one term of interest documented within their EHR notes.

Classification of CU was close to human performance for the ‘true mention’ and ‘indication of use’ categories across all models, with an F-score range of 93.2 % to 95.7% and 84.2% to 88.0%, respectively (Table 2). ClinicalBERT achieved the highest performance for the ‘true mention’ and ‘indication of use’ categories with a weighted average F-score of 92.4%. Except for the ‘patient use’ category, the performance thresholds were comparatively lower for all models for the ‘medical use,’ ‘current use,’ and ‘history of use’ categories. For these remaining categories, SVM performed slightly better than ClinicalBERT (weighted average F-score 81.6% vs. F-score 80.8%). SVM also performed the best for identifying medical use (P-82.8%, R-58.7%, F-68.4%) and current use (P-78.1%, R-68.1%, F-72.7%), while ClinicalBERT performed the best for identifying patient use (P-95.4%, R-95.1%, F-95.3%) and past use (P-57.9%, R-74.1%, F-64.5%) (Table 3).

**Table 2.** Performance metrics of algorithms for identifying cannabis users from preprocessed Electronic Health Record notes data from Geisinger. The models were run on a randomly selected and manually annotated sample of 3,650 notes. BERT: Bidirectional Encoder Representations from Transformers; SVM: Support Vector Machine.

|  |  | **Logistic**<br>**Regression** | **BERT** | **SVM** | **Clinical**<br>**BERT** |
| --- | --- | --- | --- | --- | --- |
| **True Mention**<br>**(n=1945)** | *Precision* | 0.95 | 0.91 | 0.97 | 0.96 |
|  | *Recall* | 0.96 | 0.96 | 0.94 | 0.96 |
|  | *F1 Score* | 0.95 | 0.93 | 0.96 | 0.96 |
| **Indication of Use**<br>**(n=1496)** | *Precision* | 0.85 | 0.81 | 0.87 | 0.86 |
|  | *Recall* | 0.88 | 0.88 | 0.86 | 0.90 |
|  | *F1 Score* | 0.87 | 0.84 | 0.86 | 0.88 |
| **Weighted Average** | *Precision* | 0.90 | 0.86 | 0.93 | 0.91 |
|  | *Recall* | 0.92 | 0.92 | 0.91 | 0.93 |
|  | *F1 score* | 0.91 | 0.89 | 0.92 | 0.92 |

**Table 3.** Performance metrics of algorithms for identifying characteristics of cannabis use context documented in Geisinger Electronic Health Record notes. The models were run on a randomly selected and manually annotated sample of 3650 notes. BERT: Bidirectional Encoder Representations from Transformers; SVM: Support Vector Machine.

|  |  | **Logistic**<br>**Regression** | **BERT** | **SVM** | **Clinical**<br>**BERT** |
| --- | --- | --- | --- | --- | --- |
| **Patient Use**<br>**(n=1,911)** | Precision | 0.94 | 0.91 | 0.96 | 0.95 |
|  | Recall | 0.95 | 0.94 | 0.94 | 0.95 |
|  | F1 Score | 0.94 | 0.92 | 0.95 | 0.95 |
| **Medical Use**<br>**(n=359)** | Precision | 0.82 | 0.58 | 0.83 | 0.52 |
|  | Recall | 0.33 | 0.71 | 0.59 | 0.86 |
|  | F1 Score | 0.46 | 0.61 | 0.68 | 0.65 |
| **Current Use**<br>**(n=973)** | Precision | 0.76 | 0.60 | 0.78 | 0.60 |
|  | Recall | 0.60 | 0.74 | 0.68 | 0.80 |
|  | F1 Score | 0.67 | 0.66 | 0.73 | 0.68 |
| **Past Use (n=551)** | Precision | 0.73 | 0.61 | 0.74 | 0.58 |
|  | Recall | 0.41 | 0.69 | 0.51 | 0.74 |
|  | F1 Score | 0.52 | 0.64 | 0.60 | 0.65 |
| **Weighted Average** | Precision | 0.85 | 0.75 | 0.87 | 0.77 |
|  | Recall | 0.72 | 0.83 | 0.78 | 0.87 |
|  | F1 Score | 0.77 | 0.79 | 0.82 | 0.81 |

The trained ClinicalBERT models for ‘True Mention’ and ‘Indication of use’ subsequently classified over two-fifths (40.6%, 150,726) of the patients from the analytical sample as positive for CU. This represented 8.6% of the overall population. The distributions of all demographic variables were statistically different between the annotated sample, CU documented population, and the overall population (p < 0.0001). The average age of patients with CU documentation was similar to that of the overall population (CU - 37.6 (SD – 18.8) vs. overall - 38.1 (SD-24.6)). The proportion of patients over 65 (CU-9.3% vs. overall-17%) and under 18 (CU-11.1% vs. overall-24.6%) was higher in the overall population than in patients with CU documented in their EHR. Additionally, just over half of patients with CU were male (52.5%), compared with the overall population, where just over half were female (52.3%). Patients with CU were less likely to be Asian and more likely to be African American than the overall population. Nearly two-thirds of cannabis users (62.8%) had a Body Mass Index (BMI) greater than 25, and of these, 36.8% had a BMI ≥ 30 (i.e., obese). The proportion of obese patients was lower in the overall population (23.9%). The average BMI of cannabis users was also greater than the overall population (CU mean= 28.5, SD=8.6 vs. overall mean= 26.9, SD=9.0, Table 4). The distribution of some patient characteristics also differed between the annotated sample and those for CU and the overall population, as the random sample used for annotation was based on CU-related terminology of interest. (Table 4). Upon comparing substance use behaviors between cannabis users and overall population, we observed that CU were unlike the overall population in their current or history of use of various drugs in that they were ten-fold more likely to report tobacco smoking (49.3 % vs. 5.1%), ten-fold more likely to report alcohol use (48.2% vs. 4.9%), and nine-fold more likely to report illicit drug use (4.7% vs. 0.5%, Fig. 2).

**Table 4.** Breakdown of demographic characteristics in Geisinger patients from the annotated sample of 3173 unique patients, patients positively classified as cannabis users (CU) from a total analytical sample of 370,885 unique patients, and the overall population of more than 1.7 million unique patient at the time of either first cannabis use documentation, or first encounter documented within the electronic health records (EHR) during the study period (1/2013-6/2022). SD: Standard Deviation.

| **Total (n) –** | **Annotated**<br>**Sample** | **Cannabis Use in EHR** | **Overall population** |
| --- | --- | --- | --- |
| **Total patient (N)** | 3,173 | 150,726 | 1,762,548 |
| **Demographics\*\*** |  |  |  |
| **Age, in years** |  |  |  |
| **Mean (SD)** | 40.2 (21.8) | 37.6 (18.8) | 38.1 (24.6) |
| **< 18, N (%)** | 534 (16.8%) | 16,786 (11.1%) | 432,621 (24.6%) |
| **18-30, N (%)** | 640 (20.2%) | 46,324 (30.7%) | 314,511 (17.8%) |
| **31-45, N (%)** | 640 (20.2%) | 36,395 (24.1%) | 297,333 (16.9%) |
| **46-64, N (%)** | 897 (21.8%) | 37,276 (24.7%) | 418,268 (23.7%) |
| **65+, N (%)** | 462 (14.5%) | 14,045 (9.3%) | 299,815 (17.0%) |
| **Sex, N (%)** |  |  |  |
| **Male** | 1534 (48.4%) | 79,180 (52.5%) | 840,193 (47.7%) |
| **Female** | 1639 (52.7%) | 71,534 (47.5%) | 922,199 (52.3%) |
| **Race, <sup>a</sup> N (%)** |  |  |  |
| **White** | 2884 (90.9%) | 132,942 (88.2%) | 1,587,810 (90.1%) |
| **African American** | 208 (6.6%) | 13,896 (9.2%) | 100,816 (5.7%) |
| **Asian** | 25 (0.8%) | 687 (0.5%) | 25,961 (1.5%) |
| **Other** | 56 (1.8%) | 3,201 (2.5%) | 47,961 (2.7%) |
| **Ethnicity, N (%)** |  |  |  |
| **Non-Hispanic** | 2929 (92.3%) | 139,157 (92.3%) | 1,617,7259 (91.8%) |
| **Hispanic** | 216 (6.8%) | 9,835 (6.5%) | 112,801 (6.4%) |
| **Type of other insurance at the most recent encounter, N members (%)** |  |  |  |
| **Geisinger Health Plan** | 1122 (35.4%) | 49,240 (32.7%) | 439,333 (24.9%) |
| **Commercial** | 679 (21.4%) | 138,008 (25.2%) | 645,661 (36.6%) |
| **Medicare** | 702 (22.2%) | 21,420 (14.2%) | 321,307 (18.2%) |
| **Medicaid** | 438 (13.8%) | 24,585 (16.3%) | 141,901 (8.1%) |
| **Self-pay** | 161 (5.1%) | 13,573 (9.0%) | 158,200 (9.0%) |
| **Other/Unknown** | 71 (2.2%) | 3,900 (2.6%) | 56,146 (3.2%) |
| **BMI at index date** |  |  |  |
| **Mean (SD)** | 28.9 (9.1) | 28.5 (8.6) | 26.9 (9.0) |
| **Median (IQR)** | 28 (10.4) | 27.3 (10.2) | 26.4 (10.8) |
| **<25.0, N (%)** | 1000 (32.9%) | 49,272 (37.2%) | 539,818 (30.6%) |
| **25.0-29.9 N (%)** | 794 (26.1%) | 34,503 (26.0%) | 316,728 (18.0%) |
| **>= 30.0 – 34.9, N (%)** | 615 (20.2%) | 23,132 (17.4%) | 215,337 (12.2%) |
| **>35.0, N (%)** | 630 (20.7%) | 25,734 (19.4%) | 206,634 (11.7%) |

<sup>a</sup> - Patients could select multiple racial categories at the time of self-reporting.
\*\* p-value < 0.0001

**Fig. 2.**
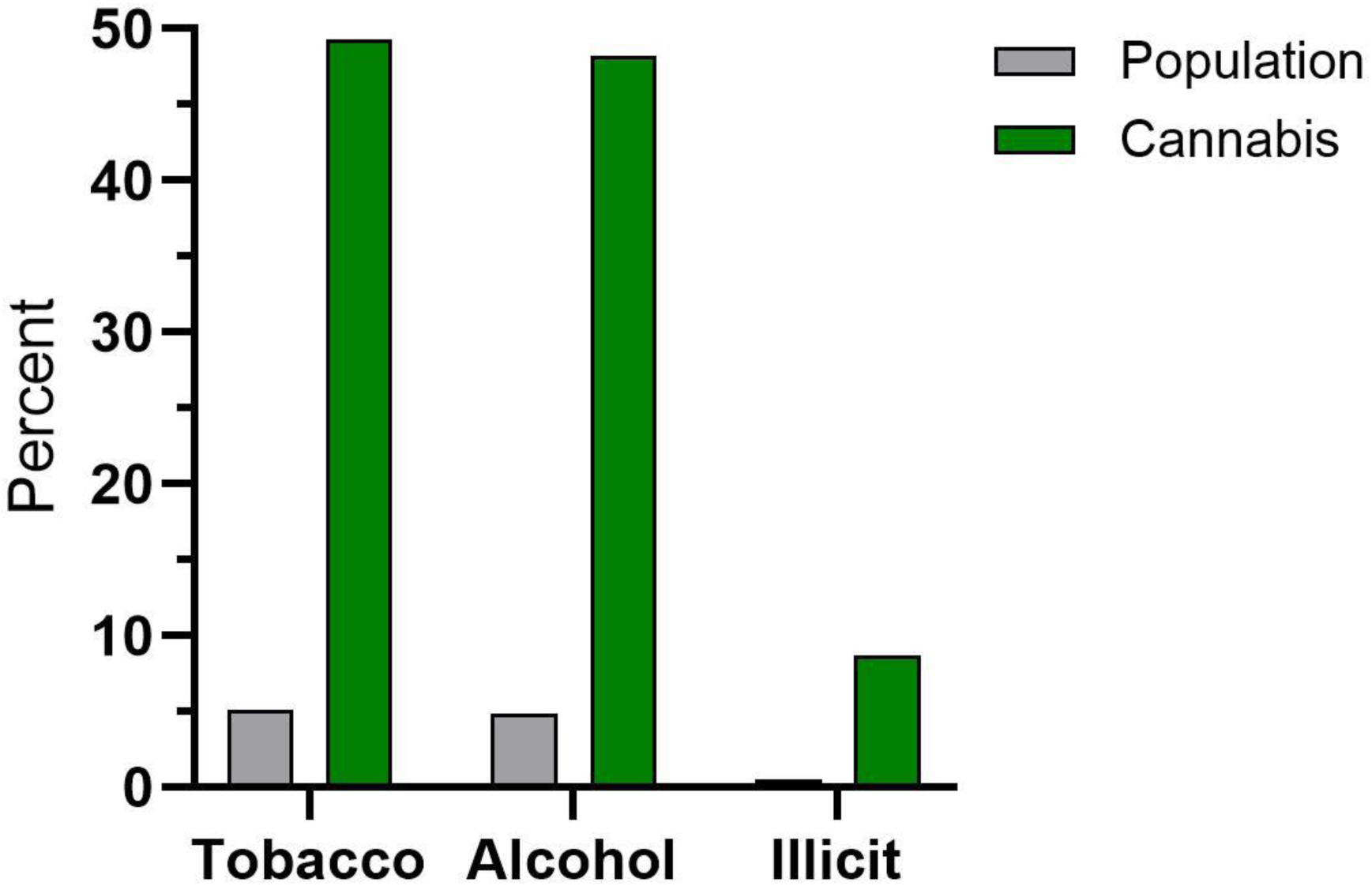
Increased prevalence of smoking tobacco (9.7 fold), drinking alcohol (9.8 fold), and illicit substance use (8.7 fold) among cannabis users, relative to the overall population, identified with natural language processing of electronic medical records among Geisinger patients seen between 2013 and 2022.

## Discussion

This study used NLP algorithms to identify patients within an integrated health system over a ten-year period who had CU documented in their EHR notes. This novel NLP application builds on prior reports [26–32] and demonstrates that CU can be identified with high precision and recall from unstructured data sources using advanced NLP models. While individual model performance varied across cannabis-related categories, the models generally performed well. ClinicalBERT, a transformer-based model, and a more traditional machine learning model, the SVM, performed nearly equally well for CU identification. Despite variations in each model’s performance for the other classification tasks, such as medical, past, current, and patient use, ClinicalBERT and SVM consistently outperformed BERT and LR. Based on these models, the prevalence of CU-related documentation was 8.6% in the overall cohort curated using only the keyword search strategy.

EHR notes and other non-discrete fields are a highly sought option for documenting cannabis and non-allopathic substances and medicines, thereby making them a key source of information for understanding utilization patterns [27,41–43] [27,43–45]. The current analysis provides the first NLP-driven, overarching insight into unsegmented EHR-based unstructured notes data on CU documentation within a single health system. This study also includes a diverse cross-section of racial breakdown in rural (e.g., Sullivan County, population 5,845) and urban (e.g., Luzerne County, population 326,496) populations (Supplemental Figure 2D). The use of traditional and transformer-based NLP models to identify CU documentation within different types of EHR notes and other unstructured data sources is rapidly increasing [22,23,26,29,44,45] and models like ours could be applied and adopted across different health systems. This investigation’s rigorous manual annotation phase, with a high interrater reliability (kappa >.83), also ensured consistency in the classification of CU. Furthermore, because the models were trained on various EHR notes spanning different specialties and disease states, as well as documenting providers, the algorithms could be easily applied to further targeted clinical and epidemiological research across the system and across different disease states.

A key report from an integrated health system in Washington state determined that for every person with past-year marijuana use documented in their EHR, seven others used marijuana, but who would only disclose this on a survey [20]. CU identified in our study over 9.5 years (8.6%) was less than half that reported in PA in the 2021-2022 National Survey of Drug Use and Health among those aged ≥ 12 for marijuana use in the past year (19.2%) [46] affirming similar incomplete documentation as noted in Washington State. Despite the recent push for cannabis being reclassified as a Schedule III substance [47] and potentially soon to be reported through state’s prescription drug monitoring program, self-reported CU information will continue to be recorded in the EHR. This is because medical use accounts for only a small portion of the total use among high schoolers [1,2] as well as adults [20], and only 24 states, three Territories, and Washington, D.C. have legalized adult use, thereby resulting in a continued reliance on self-reported CU information for assessing prevalence [5,48]. A hesitancy to disclose CU is perhaps understandable as patients might perceive that this documentation could negatively impact one’s ability to possess a firearm [49], employment [50], or even child custody [51]. The authors of an EHR report in California from 2013 to 2017 (i.e., before adult use passage) of primary care patients interpreted their social history CU documentation (0.38%) as “surprisingly low” [52] and at least ten-fold below what might be anticipated based on past-year use. Even within this context of persistent cannabis stigma [53] and under-reporting of tobacco [52] and alcohol use [Supplemental Figure 3, 4], the pronounced higher rates of tobacco, alcohol, and other illicit drug use among CU relative to the overall population are clear. The higher prevalence of CU tobacco, alcohol and other illicit drug users aligns with findings from literature[54,55], There is also some evidence that indicates that CU is more likely to be documented in the EHR for substance users compared to non-users, indicating potential biased approach towards not just who is asked or tested for CU, but also when it is documented.[29]. We could potentially hypothesize that CU is higher among those who report illicit drug use, in part, because individuals who already disclose illicit drug use may feel less need to hide or underreport their CU. However, more research is needed to validate this assumption.

Demographics and clinical characteristics differed between our annotated sample, the overall population, and CU patients. However, the clinical interpretation of these findings may not be clear due to potential confounding, given the large sample sizes, and focus on having an equitable distribution of CU-related terminologies in the annotated sample over patient characteristics [56]. For example, in our study, we found a higher proportion of obese patients in the CU cohort, which is in contrast with other studies, which found that compared to non-users, the prevalence of obesity and being overweight was lower among cannabis users in the US compared to non-users due to the increase in metabolic rates and consequent reduction in BMIs [57–60]. However, in our analysis, we did not stratify patients based on length and frequency of CU, nor did we adjust for the higher prevalence of alcohol intake in this group, or potential clinical comorbidities that may be associated with high obesity prevalence, all of which may have influenced this difference in prevalence observed [59,61]. Furthermore, we observed a higher prevalence of CU documentation within African Americans and a lower prevalence amongst Asians. While these findings align with prior research, there is existing evidence that indicates that preconceived biases, stigma, and racial profiling and discrimination surrounding who is asked about or tested for CU may be a reason behind the higher prevalence observed[29,62].

The EHR landscape is rapidly evolving, with the diffusion of agentic and generative AI-based tools implemented across multiple platforms to collect and transform data, predict potential health outcomes, and improve documentation. As the landscape continues to evolve, healthcare systems will need to develop internal capacity to tailor and fine-tune universal models to meet local and system-specific needs. In the context of CU, models developed as part of this study raise the potential for this technology to improve the current capture of CU within EHR systems and to fine-tune newer models in the future. However, it is also noteworthy that the NLP models in the current analysis performed poorly while classifying medical use of cannabis, with F1 scores ranging from 45% to 68%, indicating that the algorithms accurately classified medical use only about half the time. The distinction between medical use and non-medical use is legally clear but may have limited epidemiological utility. Among primary care patients in Washington state who indicated cannabis use, 40.1% categorized their use as medical, 31.8% as nonmedical, and 28.2% as both medical and non-medical [20]. Similarly, among medical patients in New England surveyed before the passage of laws for legalizing medical use, about half described their use as on a continuum that involved at least some recreational use [7, Supplemental Figure 4][17]. Among adults living in a state that condones medical use, the most common source of cannabis (57.5%) was from a non-dispensary source [63]. Together, these findings indicate that the distinction between medical and recreational CU is not clear.

Our investigation had several limitations. First, the results from this study are based on data collected from a single health system. CU documentation within a system could be influenced by the legalization status of cannabis at the time of data collection (i.e., currently condoning medical but not recreational in PA) [64], health system-level regulations, equity concerns about cannabis documentation attributable to racial diversity (Supplemental Figure 2D [29,65], and clinician-level characteristics, including but not limited to personal biases. Second, while using fivefold cross-validation improved the internal validity of these models, they were not externally validated, thereby limiting their generalizability for use at external institutions. Third, we identified the EHR notes sample using a keyword search for terminologies. There is still a possibility that some terminologies, variations, or misspellings could have been missed (e.g., “marijuana” can be misspelled as “marihuana” or “marjiuana” or “marajuana”), impacting the number of notes identified and restricting the sample. We curated literature and sought stakeholder input to assemble a comprehensive lexicon that would minimize the impact of this limitation. Additionally, based on our models, we could not conclude medical use or distinguish between current and past use with certainty due to poor model performance. This could be attributed to semantic variability in the documentation of this information and to low coder agreement during the manual annotation phase when classifying these factors. Prior research has encountered similar difficulties when leveraging NLP algorithms [26,27]. Consistency in documenting practices across the system may help address ambiguity in reported information and improve model performance. Lastly, data captured in the EHR remains susceptible to the general limitations of EHR data, such as inaccuracy, incompleteness, and insufficient granularity [66]. System-level initiatives that ensure regular data quality checks could help overcome this.

## Conclusion

Our study successfully leveraged NLP methodologies to classify CU documentation from EHR notes with high precision. System-level initiatives supporting automated extraction of this information using NLP and its potential transformation into data that can be discretely queried, as well as leveraging the rapidly evolving artificial intelligence capabilities within the EHR, may lead to an improved understanding of patients’ health and facilitate future clinical and epidemiological research surrounding CU prevalence and patterns among different patient populations.

## Supporting information

This file contains four supplemental figures and Supplements A-F

## Acknowledgement

We would like to thank our Summer Research Immersion Program student, Ms. Alivia Roberts, and Ms. Faith Garasich, a Summer Undergraduate Research Program student, for their assistance in the manual annotation work on this study, and Wei Wei, PhD, and Joseph J. Dewalle, for fruitful discussions, as well as Megan Gunther and Iris Johnston for technical support. Vanessa Troiani is currently the director of the Geisinger’s Academic Clinical Research Center, which operates through the Center for Substance Use Research and Education.

## Statement of Ethics

This study protocol was reviewed and approved by the Geisinger IRB (approval number *2022-0498)*.

## Author Contributions

All authors contributed to the study design and conceptualization of the manuscript. AMP, BJP, and VS drafted the initial manuscript. CMG and AMP drafted the supplemental material. AMP developed the initial lexicon that was revised, edited, and approved by JG, BJP, VS, and EAW. AMP, VS, and JG performed the data preprocessing. AMP and JG performed the data extraction. VS drafted the initial annotation guidelines, which were revised, approved, and finalized by AMP, BJP, LT, EAW, and CMG. CMG, AMP, and VS performed manual annotation. VS performed inter-rater reliability analysis. AMP and VS performed the final data analysis. AMP, VS, CMG, LT, BJP, JG, and EW revised and approved the results and conclusions. All authors edited, revised, and approved the final manuscript.

## Data Availability Statement

Data and models may be shared upon reasonable request in the future, contingent on IRB approval. Specific requests for access to the data should be directed to the corresponding author, who will facilitate the process, contingent upon approval and adherence to the necessary ethical and institutional guidelines.

## References

1. Miech RA, Johnston LD, Patrick ME, O’Malley PM. Monitoring the Future national survey results on drug use, 1975-2024: overview and key findings for secondary school students. Monitoring the Future Monograph Series [Internet]. Ann Arbor, MI: Institute for Social Research, University of Michigan; 2025 [cited 2025 Mar 12]. Available from: https://monitoringthefuture.org/results/annual-reports/.

2. Miech RA, Johnston LD, Patrick ME, O’Malley PM, Bachman JG, Schulenberg JE. Monitoring the Future national survey results on drug use, 1975-2022: secondary school Students [Internet]. Ann Arbor, MI: Institute for Social Research, University of Michigan; 2023 [cited 2026 Jan 23]. Available from: https://monitoringthefuture.org/results/publications/monographs/.

3. Substance Abuse and Mental Health Services Administration. Key substance use and mental health indicators in the United States: results from the 2022 National Survey on Drug Use and Health (HHS Publication No. PEP23-07-01-006, NSDUH Series H-58) [Internet]. Rockville, MD: Center for Behavioral Health Statistics and Quality, Substance Abuse and Mental Health Services Administration; 2023 [cited 2025 Mar 19]. Available from: https://www.samhsa.gov/data/report/2022-nsduh-annual-national-report.

4. Substance Abuse and Mental Health Services Administration. Key substance use and mental health indicators in the United States: results from the 2024 National Survey on Drug Use and Health (HHS Publication No. PEP25-07-007, NSDUH Series H-60) [Internet]. Rockville, MD: Center for Behavioral Health Statistics and Quality, Substance Abuse and Mental Health Services Administration; 2025 [cited 2025 Dec 23]. Available from: https://www.samhsa.gov/data/sites/default/files/reports/rpt56287/2024-nsduh-annual-national-report.pdf.

5. Hirsch AG, Wright EA, Nordberg CM, DeWalle J, Stains EL, Kennalley AL, et al. Dispensaries and medical marijuana certifications and indications: unveiling the geographic connections in Pennsylvania, USA. Med Cannabis Cannabinoids. 2024;7(1):34–43.

6. Pennsylvania Department of Health. [Internet]. Medical marijuana advisory board meeting [cited 2025 Dec 23]. Available from: https://www.pa.gov/content/dam/copapwp-pagov/en/health/documents/topics/documents/programs/medical-marijuana/Nov%2019_2025_MMAB%20Mtg.pdf.

7. Mahon E. [Internet]. A behind-the-scenes look at Spotlight PA’s analysis of 1M medical marijuana certifications [cited 2025 Mar 12]. Available from: https://www.pennlive.com/news/2023/01/a-behind-the-scenes-look-at-spotlight-pas-analysis-of-1m-medical-marijuana-certifications.html.

8. Pennsylvania Department of Health. [Internet]. Pennsylvania Medical Marijuana Program [cited 2026 Feb 13]. Available from: https://www.pa.gov/agencies/health/programs/medical-marijuana.

9. Mahon E. [Internet]. Health officials in PA face scrutiny over weak oversight of medical marijuana doctors [cited 2025 Mar 12]. Available from: https://www.spotlightpa.org/news/2022/12/pa-medical-marijuana-cards-telemedicine-doctor-oversight/.

10. Reed MK, Kelly EL, Wagner B, Hajjar E, Garber G, Worster B. A failure to guide: patient experiences within a state-run cannabis program in Pennsylvania, United States. Subst Use Misuse. 2022;57(4):516–21.

11. Steuart SR. The addition of cannabis to prescription drug monitoring programs and medication fills in Medicaid. Health Econ. 2025;34(2):283–96.

12. Nachnani R, Knehans A, Neighbors JD, Kocis PT, Lee T, Tegeler K, et al. Systematic review of drug-drug interactions of delta-9-tetrahydrocannabinol, cannabidiol, and cannabis. Front Pharmacol. 2024;15:1282831.

13. Ahrens E, Wachtendorf LJ, Hill KP, Schaefer MS. Considerations for anesthesia in older adults with cannabis use. Drugs Aging. 2024;40(12):933–43.

14. National Academies of Sciences, Engineering, and Medicine Committee on the Health Effects of Marijuana: An Evidence Review and Research Agenda Board on Population Health and Public Health Practice Health and Medicine Division. The health effects of cannabis and cannabinoids: the current state of evidence and recommendations for research. 1st ed. Washington, DC: National Academies Press; 2017.

15. Stains EL, Kennalley AL, Tian M, Boehnke KF, Kraus CK, Piper BJ. Medical cannabis in the United States: comparing 2017 and 2024 state qualifying conditions to the 2017 National Academies of Sciences report. Mayo Clin Proc Innov Qual Outcomes. 2025;9(2):100590.

16. Fearby N, Penman S, Thanos P. Effects of Δ9-tetrahydrocannibinol (THC) on obesity at different stages of life: a literature review. Int J Environ Res Public Health. 2022;19(6):3174.

17. Piper BJ, DeKeuster RM, Beals ML, Cobb CM, Burchman CA, Perkinson L, et al. Substitution of medical cannabis for pharmaceutical agents for pain, anxiety, and sleep. J Psychopharmacol. 2017;31(5):569–75.

18. Smith SA, Safwat MA, Piper BJ, Addison MA. Unraveling the enigma of cannabinoid hyperemesis syndrome: a narrative review of diagnosis and management. Cureus. 2025;17(8):e90961.

19. Hammig B, Jones C, Haldeman S. Pediatric poisonings associated with ingestion of marijuana products. J Emerg Med. 2023;64(2):181–5.

20. Lapham GT, Matson TE, Carrell DS, Bobb JF, Luce C, Oliver MM, et al. Comparison of medical cannabis use reported on a confidential survey vs documented in the electronic health record among primary care patients. JAMA Netw Open. 2022;5(5):e2211677.

21. Gregor CM, Tian MY, Tusing LD, Wright EA, Piper BJ, Romagnoli KM. State policies and clinicians’ and administrators’ perspectives on the inclusion of medical cannabis information in the Prescription Drug Monitoring Program. medRxiv [Preprint]. 2025 [cited 2026-01-22]. doi: 10.1101/2025.07.24.25332003.

22. Campbell A, Bailey SR, Hoffman KA, Ponce-Terashima J, Fankhauser K, Marino M, et al. Associations between psychiatric disorders and cannabis-related disorders documented in electronic health records. J.Psychoactive Drugs. 2020;52(3):228–36.

23. Javanbakht M, Takada S, Akabike W, Shoptaw S, Gelberg L. Cannabis use, comorbidities, and prescription medication use among older adults in a large healthcare system in Los Angeles, CA 2019–2020. J Am Geriatr Soc. 2022;70(6):1673–84.

24. Vassar M, Matthew H. The retrospective chart review: important methodological considerations. J Educ Eval Health Prof. 2013;10:12.

25. Beiler D, Chopra A, Gregor CM, Tusing LD, Pradhan AM, Romagnoli KM, et al. Medical marijuana documentation practices in patient electronic health records: retrospective observational study using smart data elements and a review of medical records. JMIR Form Res. 2024;8:e65957.

26. Sajdeya R, Mardini MT, Tighe PJ, Ison RL, Bai C, Jugl S, et al. Developing and validating a natural language processing algorithm to extract preoperative cannabis use status documentation from unstructured narrative clinical notes. J Am Med Inform Assoc. 2023;30(8):1418–28.

27. Carrell DS, Cronkite DJ, Shea M, Oliver M, Luce C, Matson TE, et al. Clinical documentation of patient-reported medical cannabis use in primary care: toward scalable extraction using natural language processing methods. Subst Abus. 2022;43(1):917–24.

28. Patel R, Wilson R, Jackson R, Ball M, Shetty H, Broadbent M, et al. Cannabis use and treatment resistance in first episode psychosis: a natural language processing study. Lancet. 2015;385 Suppl 1:S79.

29. Tavabi N, Raza M, Singh M, Golchin S, Singh H, Hogue GD, et al. Disparities in cannabis use and documentation in electronic health records among children and young adults. NPJ Digit Med. 2023;6(1):138.

30. Matson TE, Carrell DS, Bobb JF, Cronkite DJ, Oliver MM, Luce C, et al. Prevalence of medical cannabis use and associated health conditions documented in electronic health records among primary care patients in Washington State. JAMA Netw Open. 2021;4(5):e219375.

31. Sajdeya R, Rouhizadeh M, Cook RL, Ison RL, Bai C, Jugl S, et al. Cannabis use and acute postoperative pain outcomes in older adults: a propensity matched retrospective cohort study. Reg Anesth Pain Med. 2025;50(10):771–8.

32. Shetty VA, Gregor CM, Tusing LD, Pradhan AM, Romagnoli KM, Piper BJ, et al. Discussions of cannabis over patient portal secure messaging: content analysis. J Med Internet Res. 2024;26:e63311.

33. Young-Wolff KC, Sarovar V, Tucker L, Avalos LA, Alexeeff S, Conway A, et al. Trends in marijuana use among pregnant women with and without nausea and vomiting in pregnancy, 2009–2016. Drug Alcohol Depend. 2019;196:66–70.

34. von Elm E, Altman DG, Egger M, Pocock SJ, Gøtzsche PC, Vandenbroucke JP, et al. Strengthening the Reporting of Observational Studies in Epidemiology (STROBE) statement: guidelines for reporting observational studies. BMJ. 2007;335(7624):806–8.

35. Devlin J, Chang M, Lee K, Toutanova K, editors. BERT: Pre-training of deep bidirectional transformers for language understanding. Minneapolis, Minnesota: Association for Computational Linguistics; 2019. p. 4171–86 doi: 10.18653/v1/n19-1423.

36. Alsentzer E. [Internet]. Bio_ClinicalBERT [cited 2026 Jan 27]. Available from: https://huggingface.co/emilyalsentzer/Bio_ClinicalBERT.

37. Pedregosa F, Varoquaux G, Gramfort A, Michel V, Thirion B, Grisel O, et al. Scikit-learn: machine learning in Python. J Mach Learn. 2011;12:2825–30.

38. Wolf T, Debut L, Sanh V, Chaumond J, Clement Delangue, Moi A, et al. HuggingFace’s transformers: state-of-the-art natural language processing. 2020 [cited 2026 Jan 27]. doi: 10.48550/ARXIV.1910.03771.

39. Monaghan TF, Rahman SN, Agudelo CW, Wein AJ, Lazar JM, Everaert K, et al. Foundational statistical principles in medical research: sensitivity, specificity, positive predictive value, and negative predictive value. Medicina (Kaunas). 2021;57(5):503.

40. GitHub. [Internet]. GitHub [cited 2024 Apr 23]. Available from: https://github.com/login?return_to=https%3A%2F%2Fgithub.com%2Fnew.

41. Jafari E, Blackman MH, Karnes JH, Van Driest SL, Crawford DC, Choi L, et al. Using electronic health records for clinical pharmacology research: Challenges and considerations. Clin Transl Sci. 2024;17(7):e13871.

42. Zhang R, Manohar N, Arsoniadis E, Wang Y, Adam TJ, Pakhomov SV, et al. Evaluating term coverage of herbal and dietary supplements in electronic health records. AMIA Annu Symp Proc. 2015;2015:1361–70.

43. Sajdeya R, Goodin AJ, Tighe PJ. Cannabis use assessment and documentation in healthcare: priorities for closing the gap. Prev Med. 2021;153:106798.

44. Keyhani S, Vali M, Cohen B, Woodbridge A, Arenson M, Eilkhani E, et al. A search algorithm for identifying likely users and non-users of marijuana from the free text of the electronic medical record. PLoS One. 2018;13(3):e0193706.

45. Padwa H, Huang D, Mooney L, Grella CE, Urada D, Bell DS, et al. Medical conditions of primary care patients with documented cannabis use and cannabis use disorder in electronic health records: a case control study from an academic health system in a medical marijuana state. Subst Abuse Treat. 2022;17(1):36.

46. Substance Abuse and Mental Health Services Administration. State data tables and reports from the 2021-2022 NSDUH [Internet]. Rockville, MD: Center for Behavioral Health Statistics and Quality; 2023 [cited 2025 Mar 12]. Available from: https://www.samhsa.gov/data/sites/default/files/reports/rpt44484/2022-nsduh-sae-tables-percent-CSVs/2022-nsduh-sae-tables-percent.pdf.

47. Drug Enforcement Administration. [Internet]. DEA to hold hearing on the rescheduling of marijuana [cited 2025 Mar 12]. Available from: https://www.dea.gov/stories/2024/2024-11/2024-11-26/dea-hold-hearing-rescheduling-marijuana.

48. National Conference of State Legislatures. State medical cannabis laws [Internet]. National Conference of State Legislatures; 2025 [cited 2025 Feb 28]. Available from: https://www.ncsl.org/health/state-medical-cannabis-laws.

49. Wasserstein B. [Internet]. It’s illegal to possess both guns and medical marijuana in PA. This bill would change that [cited 2025 Mar 12]. Available from: https://www.wvia.org/news/pennsylvania-news/2024-02-19/its-illegal-to-possess-both-guns-and-medical-marijuana-in-pa-this-bill-would-change-that.

50. Mahon E. [Internet]. Unimpaired, unemployed [cited 2025 Mar 12]. Available from: https://www.spotlightpa.org/news/2022/09/pennsylvania-medical-marijuana-job-fired/.

51. Hangley Aronchick Segal Pudlin & Schiller. [Internet]. How does medical marijuana use affect child custody? [cited 2025 Mar 12]. Available from: https://www.hangley.com/blog/how-does-medical-marijuana-use-affect-child-custody.

52. Chen L, Quinn V, Xu L, Gould MK, Jacobsen SJ, Koebnick C, et al. The accuracy and trends of smoking history documentation in electronic medical records in a large managed care organization. Subst Use Misuse. 2013;48(9):731–42.

53. Bottorff JL, Bissell LJ, Balneaves LG, Oliffe JL, Capler NR, Buxton J. Perceptions of cannabis as a stigmatized medicine: a qualitative descriptive study. Harm Reduct J. 2013;10:2.

54. Secades-Villa R, Garcia-Rodríguez O, Jin CJ, Wang S, Blanco C. Probability and predictors of the cannabis gateway effect: a national study. Int.J.Drug Policy. 2015 Feb;26(2):135–42. doi:10.1016/j.drugpo.2014.07.011.

55. Subbaraman MS, Kerr WC. Cannabis use frequency, route of administration, and co-use with alcohol among older adults in Washington state. J.Cannabis Res. 2021 Jun 3;3(1):17–3. doi:10.1186/s42238-021-00071-3.

56. Gan W. Impact of sample size and its estimation in medical research. AJPM Focus. 2025:100451.

57. Cavalheiro EKFF, Costa AB, Salla DH, Silva MRd, Mendes TF, Silva LEd, et al. Cannabis sativa as a treatment for obesity: From anti-inflammatory indirect support to a promising metabolic re-establishment target. Cannabis Cannabinoid Res. 2022;7(2):135–51.

58. Hayatbakhsh MR, O’Callaghan MJ, Mamun AA, Williams GM, Clavarino A, Najman JM. Cannabis use and obesity and young adults. Am J Drug Alcohol Abuse. 2010;36(6):350–6.

59. Le Strat Y, Le Foll B. Obesity and cannabis use: results from 2 representative national surveys. Am J Epidemiol. 2011;174(8):929–33.

60. Clark TM, Jones JM, Hall AG, Tabner SA, Kmiec RL. Theoretical explanation for reduced body mass index and obesity rates in cannabis users. Cannabis Cannabinoid Res. 2018;3(1):259–71.

61. Robinson E, Humphreys G, Jones A. Alcohol, calories, and obesity: A rapid systematic review and meta-analysis of consumer knowledge, support, and behavioral effects of energy labeling on alcoholic drinks. Obes Rev. 2021;22(6):e13198.

62. Gaither JR, Gordon K, Crystal S, Edelman EJ, Kerns RD, Justice AC, et al. Racial disparities in discontinuation of long-term opioid therapy following illicit drug use among black and white patients. Drug Alcohol Depend. 2018 Nov 1;192:371–6. doi:10.1016/j.drugalcdep.2018.05.033.

63. Myers MG, Dowling MA, Bohnert KM. Where do adults in the United States obtain cannabis? A nationally representative study examining the relationships between sociodemographic factors, cannabis use characteristics and sources of cannabis. J Stud Alcohol Drugs. 2024;85(3):296–305.

64. Sajdeya R, Goodin AJ, Tighe PJ. Cannabis use assessment and documentation in healthcare: priorities for closing the gap. Prev.Med. 2021;153:106798.

65. Cohen S, Nielsen T, Chou JH, Hoeppner B, Koenigs KJ, Bernstein SN, et al. Disparities in maternal-infant drug testing, social work assessment, and custody at 5 hospitals. Acad Pediatr. 2023;23(6):1268–75.

66. Hersh WR, Weiner MG, Embi PJ, Logan JR, Payne PRO, Bernstam EV, et al. Caveats for the use of operational electronic health record data in comparative effectiveness research. Med Care. 2013 Aug;51(8 Suppl 3):S30–7.

