## Supplementary material for "Cannabis Use Documentation within the Electronic Health Record: A Use Case for Natural Language Processing Methods": This file contains four supplemental figures and Supplements A-F

**Supplemental Figure 1**. Certifications for medical marijuana in PA in 2021. The number of patients is in parentheses. Amyotrophic Lateral Sclerosis: ALS; IBD: Irritable Bowel Disorder; OUD: Opioid Use Disorder; PTSD: Post Traumatic Stress Disorder. Accessed 1/2/2026 at: https://www.bctv.org/2023/01/31/a-behind-the-scenes-look-at-spotlight-pas-analysis-of-1-million-medical-marijuana-certifications/

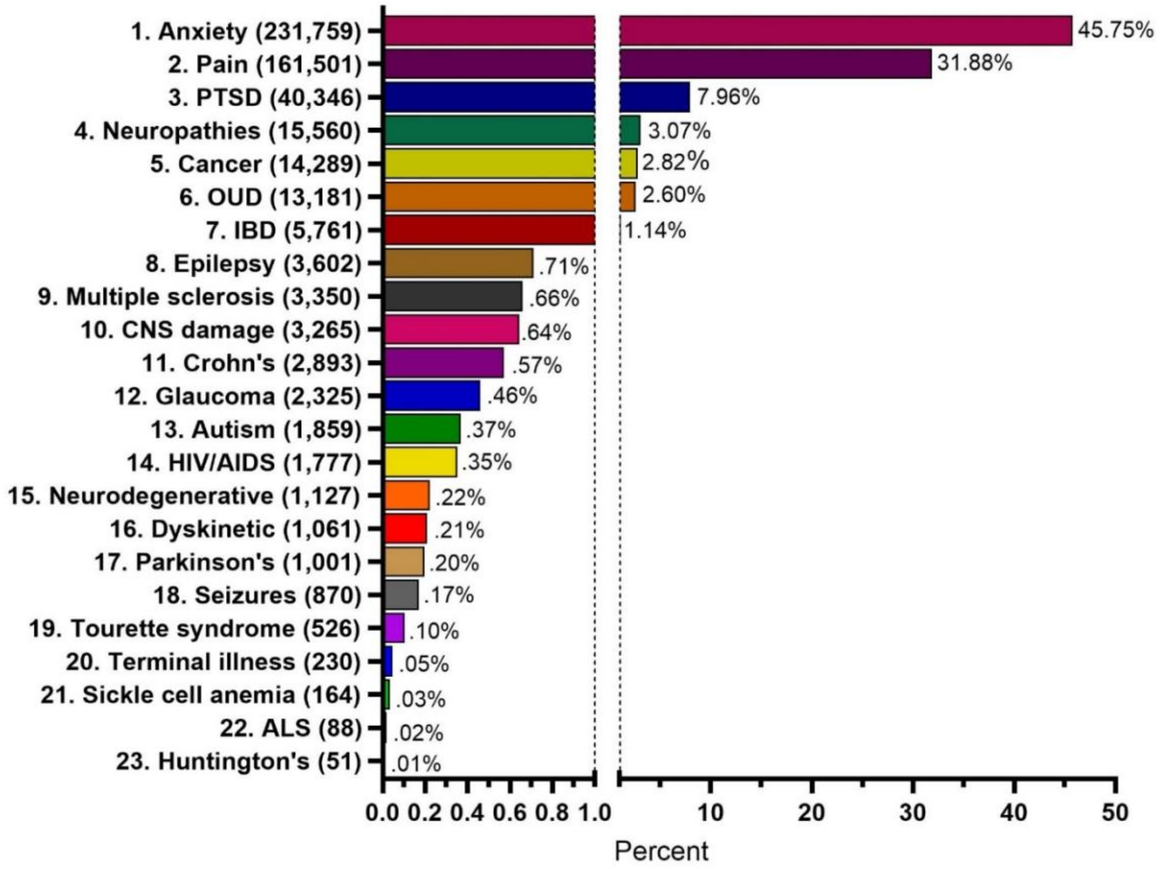

**Supplemental Figure 2A**. PA county map showing the Geisinger service area (**A**). Accessed 2/23/2025 at: https://www.geisinger.org/-/media/OneGeisinger/pdfs/ghs/about-geisinger/pdfs/Geisinger-System-Map---August-2020.pdf?sc_lang=en&hash=238C757DB7E2564CE16759D177761499

Heatmaps created with Datawrapper showing population changes from 2013 to 2022 (**2B**), the percent of the population born in that country (nativity**, 2C**), and the percent non-White (**2D**).

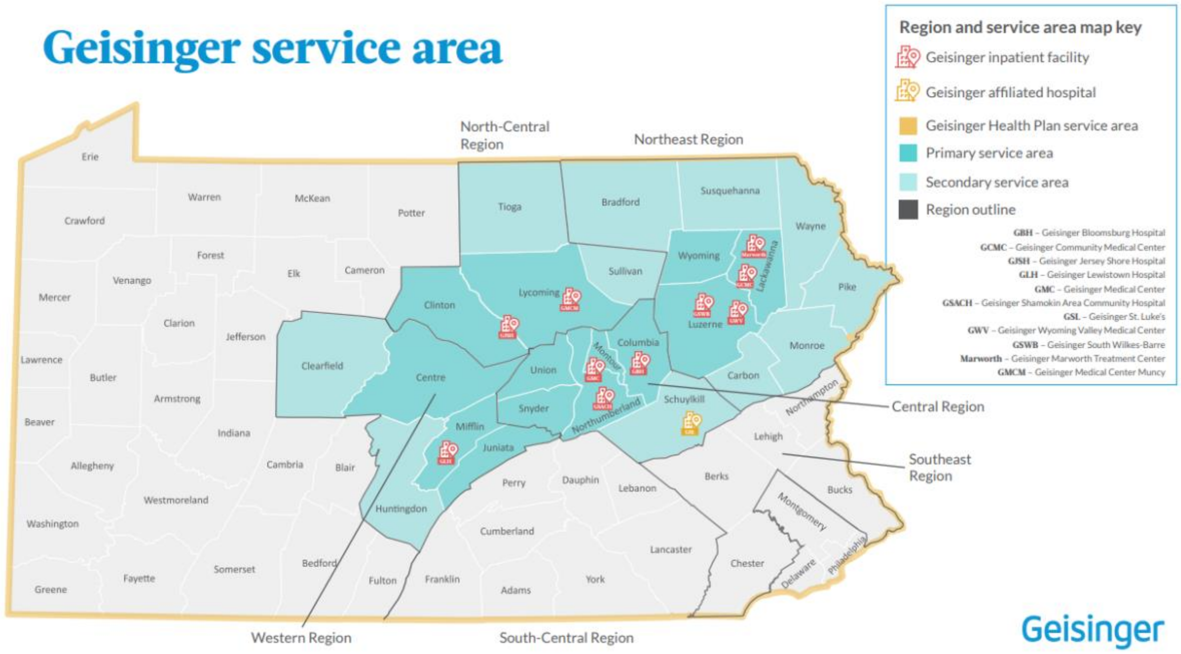

**2B.** Percent change in county population in the Geisinger primary and secondary service areas according to the American Community Survey. The population declined from 2013 to 2022 in both the primary (Mean = -2.8%, SD = 2.7%, Min = -8.0% in Wyoming, Max = +2.0% Luzerne) and secondary (Mean = -3.0%, SD = 4.1%, Min = -9.7% Susquehanna, Max = +5.6% Pike) service area counties. In comparison, PA increased +1.5%.

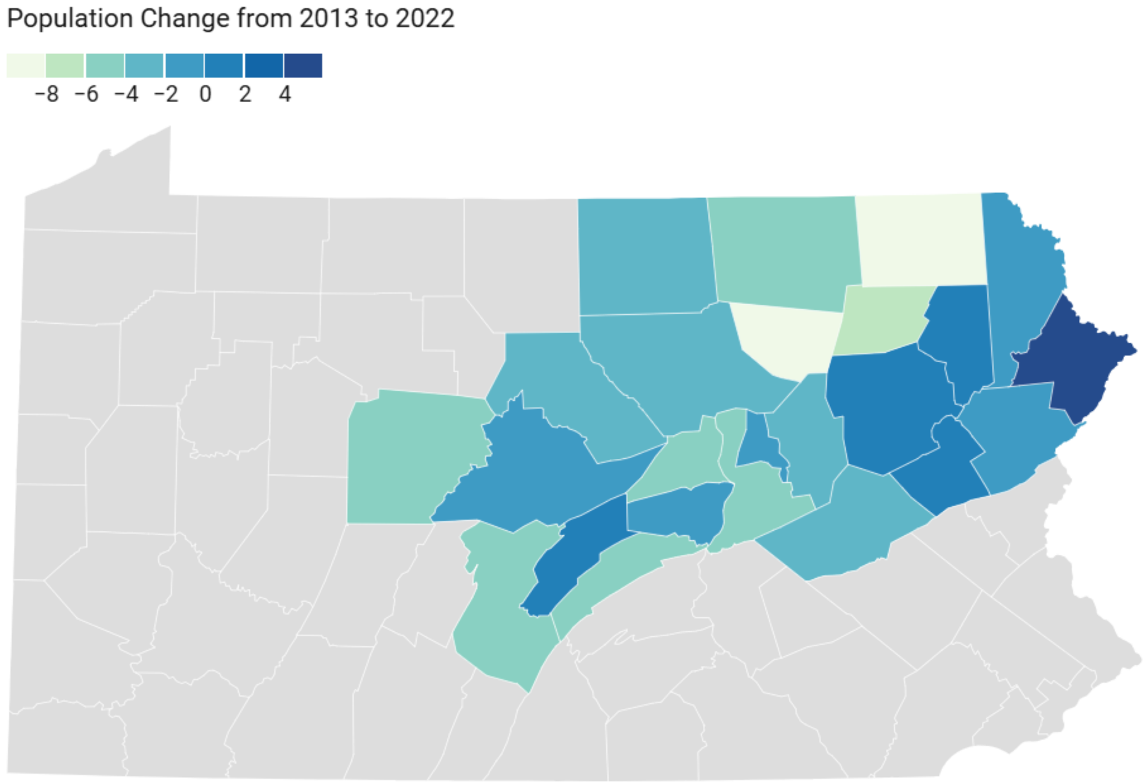

**2C.** Nativity, being born in the same county you currently live in, in 2022, according to the American Community Survey. There was high nativity in the primary (Mean = 96.7%, SD = 2.6%, Min = 91.7% in Centre, Max = 98.9% in Snyder) and secondary (Mean = 96.6%, SD = 2.8%, Min = 89.8% in Monroe, Max = 99.0% in Huntingdon) service areas relative to both PA (92.6%) and the US (86.1%).

PA source: https://www.census.gov/quickfacts/fact/table/PA/PST045224

US source: https://www2.census.gov/library/publications/2024/demo/acsbr-019.pdf

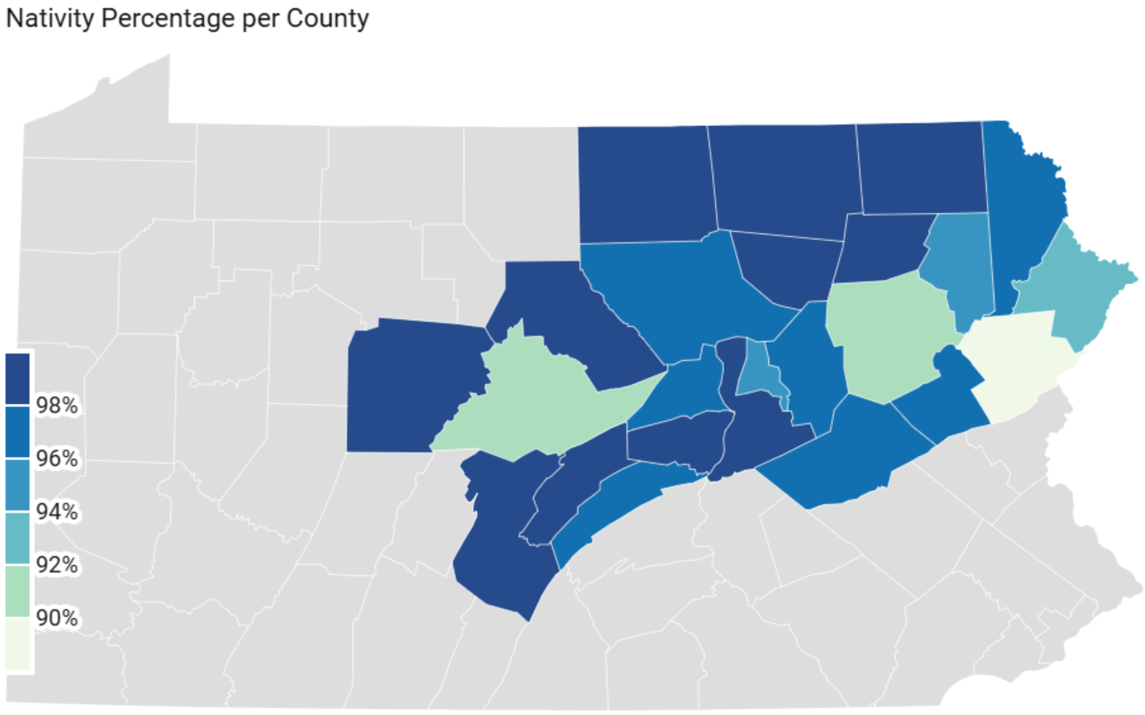

**2D**. Percent non-White population according to the American Community Survey. The percent non-White in the primary (Mean = 9.1%, SD = 4.5%, Min = 4.5% in Snyder, Max = 19.4% in Luzerne) and secondary (Mean = 10.3%, SD = 7.6%, Min = 3.7% in Susquehanna, Max = 30.6% in Monroe) service areas in 2022 was much lower than PA (19.4%) or the United States (24.7%).

Sources:

PA: https://data.census.gov/table?q=white+population+in+all+PA+counties+in+2022
United States: https://www.census.gov/quickfacts/fact/table/US/PST045224

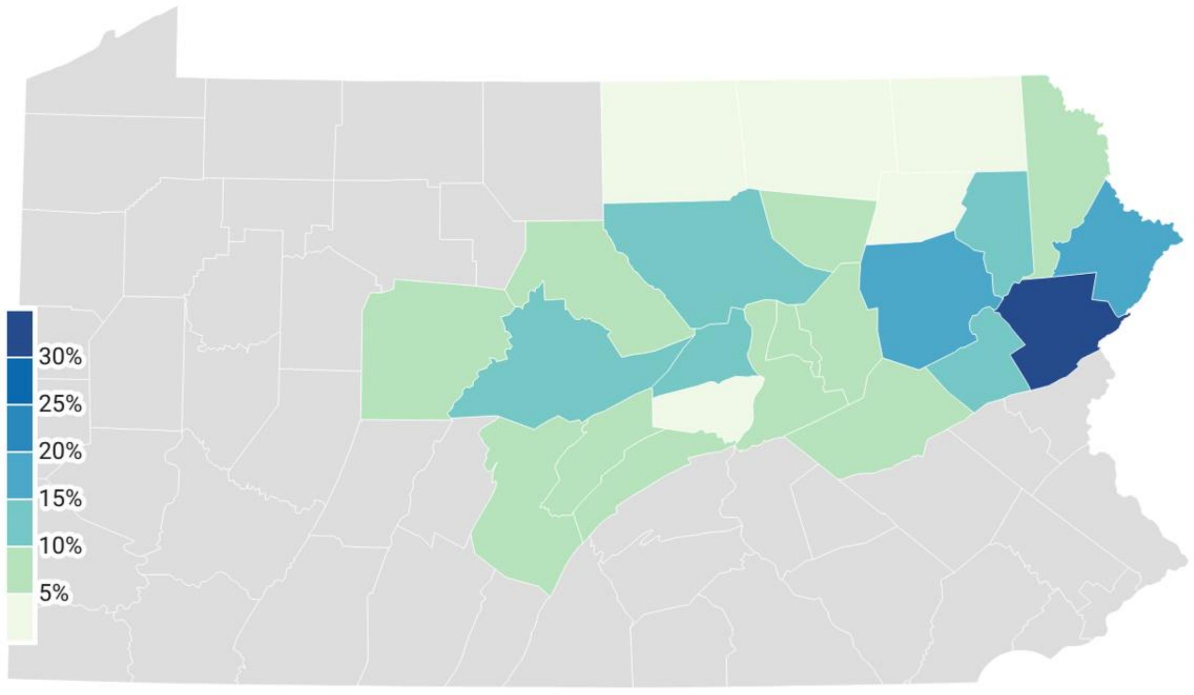

**Supplemental Figure 3.** Past-month use of various substances for those aged≥ 12 in PA in 2021 and 2022 according to the National Survey on Drug Use and Health [45].

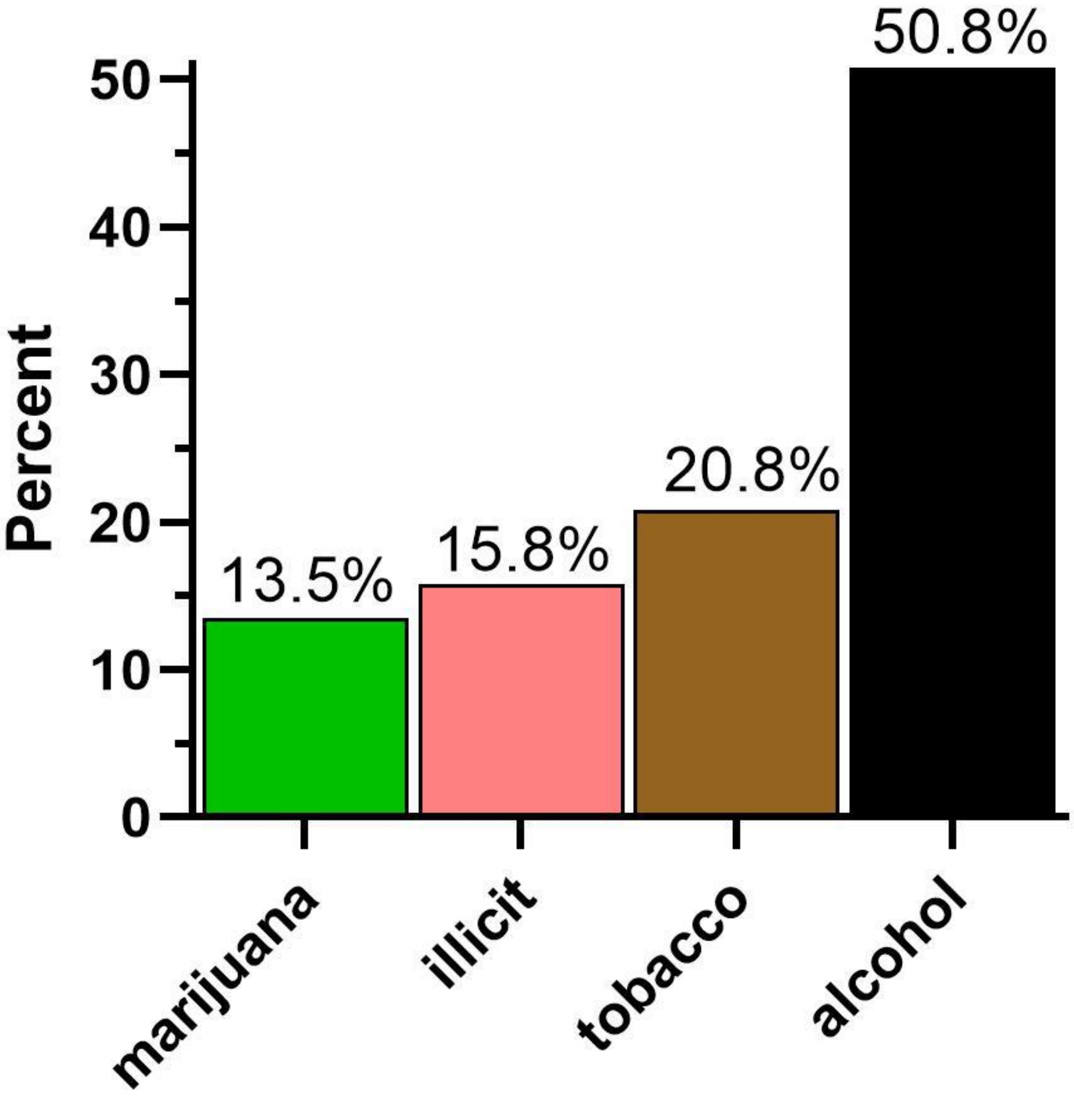

**Supplemental Figure 4.** Medical marijuana patients in Vermont (left) and Maine (right) in 2015 (i.e., before the passage of law for legalized medical use) were asked, "How would you describe your use of cannabis? For example, if you use medical cannabis twice a day for your symptoms but on the weekends, twice midday to relax with friends, you might choose 20% recreation / 80% medical." with response options ranging from 0% recreational / 100% medical to 100% recreational / 0% medical [14].

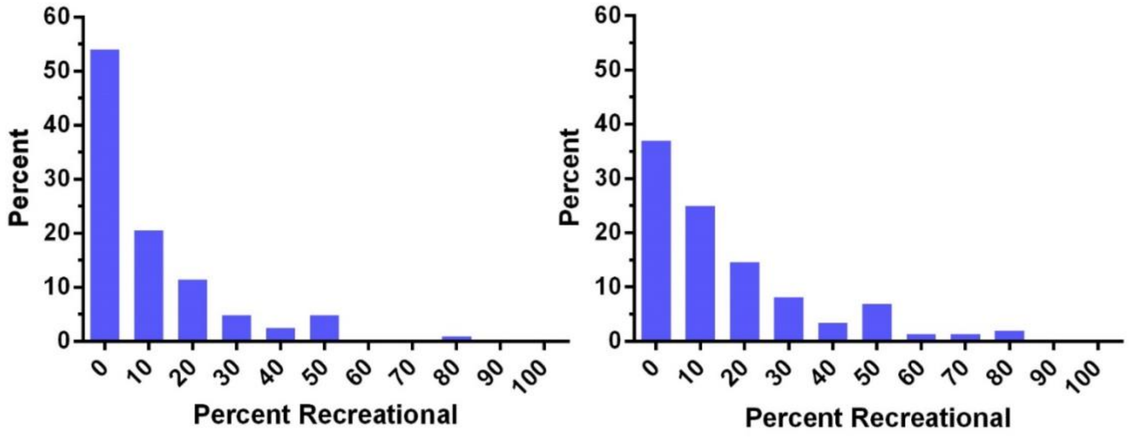

**Supplement A – Primary Data Cleaning**

**Overview**

The purpose of annotating the unstructured clinical patient notes was to train natural language processing algorithms to replicate human annotation at the mention of cannabis. We first needed to narrow our search; we developed a cannabis use (CU) lexicon to achieve this. We used an informal literature review and feedback from subject-matter experts on the study team to finalize the CU-related terms. The lexicon included the following terms: 'marijuana,' 'cannabis,' 'MJ,' 'THC,' 'CBD,' 'weed,' 'MMJ,' 'indica,' 'sativa,' 'cannabinoid,' 'spice,' 'tetrahydrocannabinol,' 'pot,' 'cannabidiol,' 'ganja,' 'grass,' 'hash,' 'hashish,' 'bong,' 'Mary Jane,' 'edibles,' 'joint.'

**Data source**

Encounter notes curated from the EHR included nursing notes, appointment notes, problem list-based free-text documentation, visit reason documentation, and follow-up notes. Nursing notes often included patients' chief complaints, medical and family histories, social histories, and other free-text documentation, such as patient assessment and plan. Pre-surgical or pre-procedural evaluations undertaken in these patients were also documented in the nursing notes. The nursing, follow-up, and appointment notes included laboratory-related orders, referrals, and documentation of medication management.

**Primary Data Cleaning**

As a first step towards cleaning the 135,858,323 EHR note files, the matching keyword search terms were capitalized, and the notes were trimmed to approximately 300 characters before and after each search term to reduce file sizes and improve readability. At a cursory glance, it was observed that multiple non-CU-related common medical phrases were being pulled in this data. To avoid misclassification, we deleted notes that included the following phrases or parts of words for each relevant term, resulting in a sample of 17,789,648 EHR notes

a. The term POT is used with "amoxicillin," "neti," or the letters POT as part of the words "hypotension," "potential," or "potassium."

b. The term SPICE with "choose ingredients that."

c. The term THC is immediately followed by the word "no."

d. The term BONG, in reference to a patient's or doctor's name

e. The term CBD is immediately followed by "pancreas," "dilation," "measure," "no," "CBD measure," and "CBD stricture."

f. The term GRASS, along with "allergy," "mowing," "mowed," "mows," "pollen," "environmental," "skin test," or "ragweed."

g. The term INDICA, but words suggesting that the note was in Spanish and being used as a verb ("se indica," "lo indica," "le indica," "indica que est")

h. The term WEED is part of the word "ragweed", or together with "pollen."

i. The term JOINT is part of the phrases "joint pain," "joint space," "joint deformity," and "joint disease."

The term ‘joint’ was also primarily and extensively used in an anatomical context, such as “joint pain,” “joint space,” “joint deformity,” and “joint disease”; therefore, notes with the term ‘joint’ were removed from further analysis. The data cleaning process also involved the removal of notes when the term of interest was negated (e.g., “no cannabidiol use” or “tetrahydrocannabinol (THC): no.”) This led to a more concise final list of terms and a final analytical sample of 2,790,896 EHR notes with at least one mention of a key term in patient charts. We then split the partially cleaned data into individual files based on the CU-related terminology they contained. A random sample was drawn from these sub-files and combined into a dataset to be shared with the annotators for assigning classification labels. The random selection did not account for any patient characteristics, but focused on representation of the CU-related term of interest.

**Supplement B: Labelling Guidelines**

The text surrounding the capitalized keyword provided context for annotators to decide whether the note was related to cannabis, and we referred to this text as snippets. There could be more than one matching keyword in the note; however, usually only one was capitalized. If there were multiple keywords, they were not analyzed separately; the whole snippet was analyzed.

An example of a note with multiple keywords:

*x95 drug use: yes types: marijuana comment: marijuana daily, history* ***HASH****, lsd, speed]*

From this note, you can see that only one of the keywords was capitalized. The snippet was analyzed as a whole rather than just the part that mentioned the past use of HASH.

There were six possible labels that a snippet could have: (A) True Mention, (B) Patient Use, (C) Indication of Use, (D) Medical Use – History, (E) Current Use, and (F) History. Annotators would assign a binary number to each label, a 1 indicating (Yes) and 0 indicating (No). Annotators used "MMJ Notes Decision Rules" to guide the annotation of the notes with predefined labels.

**Label Definitions**

**A. True Mention**

*The key term refers to cannabis, irrespective of use*

The first decision in the labeling process is whether or not the mention of cannabis in the snippet is a true mention. This mention is irrespective of use. To make this decision, the annotators decide if the mention is truly explicit, without a doubt, related to cannabis, and not related to other things, such as in reference to:

- plants or objects used with plants (e.g., flower POT, watering the GRASS, pulling the WEEDs, WEED wacker),
- food and drink holders (e.g., coffee POT, POT of tea, chicken POT pie, turmeric is a SPICE, HASH browns, Mangifera INDICA (i.e., mango) ),
- anatomy/physiology (CBD in reference to complete blood count when listing labs, CBD about common bile duct, MJ neck collar, etc.).
- energy (cumulative energy: 36.7 MJ)
- part of another word irrelevant to marijuana/cannabis drug (e.g., INDICAtes)
- part of a first, middle, or last name (e.g., a fake example of this is last name = WEED)
- cold/allergy remedy (e.g., netti POT)

- Spanish language (i.e., INDICA = indicates)

If a word was cut off and didn't have the full name, such as "TH" missing the "C" and nothing else, we evaluated the scenario on a case-by-case basis. We considered it a true mention if it made sense in the context in which the word was mentioned (e.g., Illicit Drug Use: Intermittent TH). We considered this a true mention because there is no other illicit drug that starts with TH.

If it is a true mention, the annotators record the value 1 corresponding to a true mention of cannabis (Yes). If the mention is not true, a value of 0, which is (No) cannabis, is recorded. At this point, if the decision is 0, then annotators stop annotating for this snippet and move on to the next patient note.

**B. Patient Use (Only if A = Yes)**

*True mention is about the patient*

Once true mention is determined, the annotators then decide if the mention of cannabis was related to the patient or someone other than the patient. If the snippet indicates that it refers to the patient's use, annotators record the value 1 (Yes) to indicate that the true mention is about the patient. If the mention was related to someone else's use outside the patient's use, annotators recorded 0 (No); this does not reference the patient. If the snippet was determined not to be about the patient’s use, the annotators stopped annotating for that snippet and moved on to the next patient note.

**C. Indication of Use (Only if B = Yes)**

*Explicit mention that the patient at some point in time has used marijuana/cannabis/etc.*

If the answer was 1 (Yes) to patient use, then annotators decided if the mention was an explicit mention that the patient at some point in time had used marijuana/cannabis/etc. Accidental use was not considered as having purposely used marijuana/cannabis/etc, so annotators entered 0, which was (No) the patient had not used marijuana/cannabis/etc.

Annotators had minimal medical knowledge and often could not ascertain if the patient explicitly used cannabis at some point in time; when such uncertainty arose, the snippets were reviewed by the team together, and a decision was made. If the team did not feel the mention was explicit, a 0 value was recorded, indicating No. If the decision was 0, annotators stopped annotating for that snippet and moved on to the next patient note.

**D. Medical Use – History (Only if C = Yes)**

*Explicit mention that use is for medical marijuana/cannabis/etc.*

For this section, the note had to mention medical use explicitly

- Patient has/had a medical marijuana card or

- Patient was using marijuana for relief, but there was no evidence in the note that there was a medical marijuana card or
- The mention of CBD supplements was considered medical.

For all the above, annotators entered 1, which is (Yes) the use is for medical purposes. Over-reacting was not considered a medical reason for using cannabis, and 0 was recorded.

**E. Current Use (Only if C = Yes)**

*Explicit mention that the patient currently uses marijuana/cannabis/etc.*

An explicit mention that the patient at the time of note documentation was actively using marijuana/cannabis/etc, with no indication of cessation of use (e.g., if a person said they use marijuana occasionally, then we considered it current use. Alternatively, if a person said they use marijuana once or twice a year, it was regarded as current use and history of use.

Situations that were considered currently using:

- Positive present tense verbs, adjectives, or nouns such as:
  - drug use: **yes** types: MARIJUANA
  - MJ **use**
  - MARIJUANA **occasionally**
  - CBD oil - no benefit medical MARIJUANA with cbd - **helpful**
  - THC, **confirmatory** (urine) result value ref range methodology lcmsms cannabinoids 36.5 (a) neg ng/ml
- Suspected use by the author of the note
- ICD diagnosis codes unrelated to abuse

Situations that were not considered currently using:

- If an allergy is stated, it is not regarded as current or past use.
- If medical is mentioned, but 'not yet currently using' is clarified.

**F. History of Use (Only if C = Yes)**

*Explicit mention that the patient has a history of using marijuana/cannabis/etc.*

Any mention of past use indicated a history of use (i.e., a single use in the past was considered to have a history of use as long as it was purposeful and not accidental). There wasn't a fixed look-back period assigned; it was difficult to ascertain the ideal period to consider a mention as related to past use.

The decision rule was modified to "Explicit mention" following the review of a couple of snippets that revealed that labeling the snippet for past use relied too heavily on the annotator's judgment and interpretation, resulting in significant variability in classification. The rules were updated after the review to reduce variability and improve model performance.

Examples of previously classified notes that were considered positive for a history of use before explicit mention was added to the guidelines:

- *drugs: cannabis: a couple of bowls a day. Indica at night hybrid during the day. smoking and vaping. not interested in quitting.*
- *current pain medications: medical marijuana - vape cartridges - indica at bedtime to help her sleep*
- *drug use: yes special: Marijuana comment: once in a whil*e

These snippets reveal the variability in labeling due to annotators' subjectivity in judgment and interpretation. After the criteria for past use were modified, a new annotator analyzed the examples above, and they were subsequently classified as 0 because they didn't explicitly indicate past use.

Similarly, if an ICD code was mentioned in the snippet that had no mention of abuse or disorder, such as "Marijuana use f12.90," which is for cannabis use unspecified uncomplicated, annotators did not consider this past use and recorded a 0, which is (No) the patient does not have a history of use. Section headings that included the word 'history' as a part of the title, e.g., Substance Use History:, Substance Abuse History:, Social History:, were not considered explicit enough by themselves, if there was not additional indication of past use documented along with them.

Situations that were considered history of use:

- Positive past tense verb (e.g., used)
- Negative adverb concerning time (e.g., not currently, not recently)
- The author mentions abuse or use disorder

**Supplement C: MMJ Notes Decision Rules**

**A. True Mention**

The key term refers to cannabis, irrespective of use

*No - 0*

use*s - 1*

**B. Patient Use (Only if A = Yes)**

True mention is about the patient

*No - 0*

*Yes - 1*

**C. Indication of Use (Only if B = Yes)**

Explicit mention that the patient at some point in time has used marijuana/cannabis/etc.

*No – 0*

*Yes – 1*

**D. Medical Use – History (Only if C = Yes)**

Explicit mention that use is for medical marijuana/cannabis/etc.

*No – 0*

*Yes – 1*

**E. Current Use (Only if C = Yes)**

Explicit mention that the patient currently uses marijuana/cannabis/etc.

*No – 0*

*Yes – 1*

**F. History of Use (Only if C = Yes)**

Explicit mention that the patient has a history of using marijuana/cannabis/etc.

*No – 0*

*Yes – 1*

Using the same snippet from earlier, the following is an example of labeling a note:

*x95 drug use: yes types: marijuana comment: marijuana daily, history* ***HASH****, lsd, speed]*

(Yes) this is true mention, (Yes) this is patient use, (Yes) this is indication of use, there is not enough information to determine medical, so it would be no to medical use, (Yes) this is current use, and (Yes) this is history of use.

If annotators could not decide how to label the note, the note was returned to the team for agreement on an appropriate label. Occasionally, when the questions came back to the team, the decision rules just needed some extra clarity for the annotators to make decisions; sometimes, the note section at the bottom of the decision rules was updated as required and acted as a guide on what to do when a particular situation arose (see below).

*Clarifying Rules for Guidelines:*

If allergy is stated: No for Current/Past

Mention of Abuse or Use Disorder: Yes, for Past

Suspected Use by Author: Yes for current, but No for past

Positive lab test: Yes for current

If medical mentioned but not yet using: No for current

Accidental use or 1 or 2 times: No for indication of use

The patient's indication of use is for relief, but with no medical card apparent: Yes for medical

CBD supplements are considered medical

CBD, when mentioned along with a list of lab orders such as lipo, phosphorous, bun, and creat, is talking about a complete blood count test, so – No for true mention

**Supplement D:**

1. **Examples of note snippets per label and associated frequency breakdown from training data.**

| Labels | Frequency | Example of the notes |
|---|---|---|
| True mention | 1945/3650 | 1. b'dvil, ibuprofen, motrin, aleve, naproxen). 10 days prior to surgery/procedure \xb7 stop all herbal supplements, green tea, turmeric, melatonin, cbd, THC, etc. \xb7 stop all vitamins (including vitamin e) 24 hours prior to surgery/procedure \xb7 do not consume any alcohol. \xb7 do not use medical ma'<br>2. b'est interest in place. he then spoke about how much pain he was in and that was the reason he was acting like this. he insisted he just wanted some "**EDIBLES**: marajuana" if he could just go out west he could get some and everything would be good, and couldn\'t we just prescribe him that. i explained'<br>3. b's cys is ignoring. reports she is considering beginning to drug test [patient name] when he returns from his father \'s house due to concerns that father smokes **WEED** in the home and father \'s girlfriend \'s daughter sells thc oil ("dank") out of the home. therapist cautions only beginning to test if there is reas' |
| Patient Use | 1911/3650 | 1. review of patient's allergies indicates: allergen reactions \x95 marijuana (cannabis SATIVA) hives medical marijuana \x95 tramadol tremors<br>2. b'years types: cigarettes last attempt to quit: 10/01/2013 \x95 smokeless tobacco: never used \x95 alcohol use: no \x95 drug use: no comment: **MJ** \x95 sexual activity: partners: male birth control/ protection: injection other topics concern \x95 not on file social history narrat' |
| Indication of Use | 1496/3650 | 1. b'ss tobacco: current user types: snuff \x95 tobacco comment: 1 can every day substance use topics \x95 alcohol |

| | | |
|---|---|---|
| | | use: no \x95 drug use: no types: **MARIJUANA** comment: quit smoking weed exam: bp 112/71 \| pulse 68 \| temp (src) 98.5 (tympanic) \| wt 173 lbs 9.6 oz (78.744kg) \| bmi 24.91 kg/m\xb2'<br><br>2. b'6 month recheck uses medical marijuana **EDIBLES** & vap'<br><br>3. b' pt states she believes that if she kills herself she will not go to heaven so she would not act of her actions. she reports that she recently smoked **SPICE** but has not in the past week. pt had a beer a week or so ago. she reports that she has a hx of alcohol abuse and trying to quit drinking. she st'<br><br>5. hpi comments: 6:14 pm pt presents to the ed via ems. per ems, pt overdosed on **SPICE** today. |
| Medical Use | 359/3650 | 1. Patient Use b'6 month recheck uses medical marijuana **EDIBLES** & vap'<br><br>2. review of patient's allergies indicates: allergen reactions \x95 marijuana (cannabis **SATIVA**) hives medical marijuana \x95 tramadol tremors<br><br>3. b'ased dose) and methadone bid (decreased dose). reports medical marijuana is helping and has made happier. sleeping well. not smoking marijuana, using **EDIBLES** and tinctures. still taking gabapentin tid; helping. taking flexeril bid; helping. has had a little loose stool (not true diarrhea) for pa' |
| Current Use | 973/3650 | 1. b'6 month recheck uses medical marijuana **EDIBLES** & vap'<br><br>drugs: previous abuse of drugs: yes cannabis: yes, takes 2-3 hits a few times a day, daily, has tried **EDIBLES** recently<br><br>3. b'adache from this physical therapy - she would love to go to physical therapy. current pain medications: medical marijuana - vape cartridges - **INDICA** at bedtime to help her sleeptizanidine 4mg 1/2 to 1 tablet at bedtime - |

| | | |
|---|---|---|
| | | lidocaine patches - uses them everywhere, 8 at a time sometimes gaba'<br>4. b'alcohol: no current use was a problem in the past, per patient. would drink daily denies w/d symptoms drugs: cannabis: couple of bowls a day, **INDICA** at night and hybrid during the day. smoking and vaping. not interested in quitting. methamphatamine - started in 20s, last use about 7 users'<br>5. b'ck leg pain feels like a sharp, shooting pain - constant pain is worse at night has tried heating pad without relief medical marijuana card - **EDIBLES** for sleep/pain follows with ip - scs placement trial on 4/22 what makes pain worse/increasing conditions: laying/sitting too long in one' |
| History of Use | 551/3650 | 1. b'person who he feels has a substance use problem. patient finds himself frequently crying. he also feels daily stress for which she medicates with **MARIJUANA.** he has used **MARIJUANA** for many decades. he does complain of some shortness of breath. especially when he bends over to tie his shoes.'<br>2. b'ltrasound as an outpatient in the near future. he also has a history of syncopal episodes and heart palpitations which began after he stopped smoking **SPICE** last year. he was wearing a zio ambulatory cardiac monitor for this reason (which he ripped off in the ed) and was to undergo an inpatient echoc'<br>3. b' each question to get your total score. *** maximum score is 12. intervention needed: { :5500475} drugs: previous abuse of drugs: alcohol, **SPICE** rehabilitation history: history of rehabs: no longest period of sobriety: n/a personal, family, and social history occupational' |

2. **Example snippets and annotation labels with reasoning**

| **Snippet examples** | **Annotation labels** | **Reasoning** |
|---|---|---|
| b'6 month recheck uses medical marijuana EDIBLES & vap' | A True Mention = Yes = 1<br>B Patient Use = Yes = 1<br>C Indication of Use = Yes = 1<br>D Medical Use = Yes = 1<br>E Current Use = Yes = 1<br>F History of Use = No = 0 | The only one that was not explicit enough to determine from this scenario was history of use. All others were 1. |
| 10 days prior to surgery/procedure \xb7 stop all herbal supplements, green tea, turmeric, melatonin, cbd, THC, etc. | A True Mention = Yes = 1<br>B Patient Use = Yes = 1<br>C Indication of Use = No = 0 | Yes, this is a true mention, and yes, this is referring to the patient, but there was no indication that the patient had used it at any point. |
| review of patient's allergies indicates: allergen reactions \x95 cannabis SATIVA other (please comment) found in his urine | A True Mention = Yes = 1<br>B Patient Use = Yes = 1<br>C Indication of Use = Yes = 1<br>D Medical Use = Yes = 0<br>E Current Use = Yes = 1<br>F History of Use = No = 0 | The history of use and medical use could not be determined from this scenario |
| b'keless tobacco: never used \x95 tobacco comment: pt vapes substance and sexual activity \x95 alcohol use: no \x95 drug use: not currently comment: MARIJUANA \x95 sexual activity: not on file lifestyle \x95 physical activity | A True Mention = Yes = 1<br>B Patient Use = Yes = 1<br>C Indication of Use = Yes = 1<br>D Medical Use = Yes = 0<br>E Current Use = Yes = 0<br>F History of Use = Yes = 1 | History of use is (Yes) because it states not currently, but they have used it in the past. |

| |
|---|
| days per week: not on file<br>minutes pe' |

**Supplement E**. Inter-rater reliability using Cohen's kappa calculated between the three independent coders. The scores below demonstrate the agreement of Annotator 3 with Annotators 1 & 2.

| Labeling Category | Inter-rater Reliability | |
|---|---|---|
| | Annotator 1 | Annotator 2 |
| True Mention | 0.918 | 0.918 |
| Patient Use | 0.833 | 0.833 |
| Indication of Use | 0.818 | 0.865 |
| Medical Use | 0.898 | 0.898 |
| Current Use | 0.896 | 0.896 |
| Past Use/ History of Use | 0.615 | 0.565 |

**Supplement F:** STROBE Statement—Checklist of items that should be included in reports of *cross-sectional studies*.

| | Item No | Recommendation | Page number |
|---|---|---|---|
| Title and abstract | 1 | (*a*) Indicate the study's design with a commonly used term in the title or the abstract | 1,2 |
| | | (*b*) Provide in the abstract an informative and balanced summary of what was done and what was found | |
| Introduction | | | 5,6 |
| Background/rationale | 2 | Explain the scientific background and rationale for the investigation being reported | 5,6 |
| Objectives | 3 | State specific objectives, including any prespecified hypotheses | 5,6 |
| Methods | | | 7-10 |
| Study design | 4 | Present key elements of study design early in the paper | 7 |
| Setting | 5 | Describe the setting, locations, and relevant dates, including periods of recruitment, exposure, follow-up, and data collection | 7 |
| Participants | 6 | (*a*) Give the eligibility criteria, and the sources and methods of selection of participants | 7 |
| Variables | 7 | Clearly define all outcomes, exposures, predictors, potential confounders, and effect modifiers. Give diagnostic criteria, if applicable | 8-10 |
| Data sources/ measurement | 8* | For each variable of interest, give sources of data and details of methods of assessment (measurement). Describe comparability of assessment methods if there is more than one group | 8-10 |
| Bias | 9 | Describe any efforts to address potential sources of bias | 8-10 |
| Study size | 10 | Explain how the study size was arrived at | 7-9 |

| | | | |
|---|---|---|---|
| Quantitative variables | 11 | Explain how quantitative variables were handled in the analyses. If applicable, describe which groupings were chosen and why | 8-10 |
| Statistical methods | 12 | (*a*) Describe all statistical methods, including those used to control for confounding | 9,10 |
| | | (*b*) Describe any methods used to examine subgroups and interactions | |
| | | (*c*) Explain how missing data were addressed | |
| | | (*d*) If applicable, describe analytical methods taking account of sampling strategy | |
| | | (*e*) Describe any sensitivity analyses | |
| Results | | | 11,12 |
| Participants | 13* | (a) Report numbers of individuals at each stage of study—eg numbers potentially eligible, examined for eligibility, confirmed eligible, included in the study, completing follow-up, and analysed | 11 |
| | | (b) Give reasons for non-participation at each stage | |
| | | (c) Consider use of a flow diagram | |
| Descriptive data | 14* | (a) Give characteristics of study participants (eg demographic, clinical, social) and information on exposures and potential confounders | 11,12 |
| | | (b) Indicate number of participants with missing data for each variable of interest | |
| Outcome data | 15* | Report numbers of outcome events or summary measures | 11 |
| Main results | 16 | (*a*) Give unadjusted estimates and, if applicable, confounder-adjusted estimates and their precision (eg, 95% confidence interval). Make clear which confounders were adjusted for and why they were included | 11,12 |

| | | | |
|---|---|---|---|
| | | (*b*) Report category boundaries when continuous variables were categorized | |
| | | (*c*) If relevant, consider translating estimates of relative risk into absolute risk for a meaningful time period | |
| Other analyses | 17 | Report other analyses done—eg analyses of subgroups and interactions, and sensitivity analyses | 11,12 |
| Discussion | | | 13-16 |
| Key results | 18 | Summarise key results with reference to study objectives | 13-15 |
| Limitations | 19 | Discuss limitations of the study, taking into account sources of potential bias or imprecision. Discuss both direction and magnitude of any potential bias | 15,16 |
| Interpretation | 20 | Give a cautious overall interpretation of results considering objectives, limitations, multiplicity of analyses, results from similar studies, and other relevant evidence | 15,16 |
| Generalisability | 21 | Discuss the generalisability (external validity) of the study results | 15,16 |
| Other information | | | |
| Funding | 22 | Give the source of funding and the role of the funders for the present study and, if applicable, for the original study on which the present article is based | 17 |

*Give information separately for exposed and unexposed groups.

Note: An Explanation and Elaboration article discusses each checklist item and gives methodological background and published examples of transparent reporting. The STROBE checklist is best used in conjunction with this article (freely available on the Web sites of PLoS Medicine at http://www.plosmedicine.org/, Annals of Internal Medicine at http://www.annals.org/, and Epidemiology at http://www.epidem.com/). Information on the STROBE Initiative is available at www.strobe-statement.org.
